# Pharmacokinetics-based identification of antiviral compounds of *Rheum palmatum* rhizomes and roots (Dahuang)

**DOI:** 10.1101/2023.08.28.23294750

**Authors:** Nan-Nan Tian, Ling-Ling Ren, Ya-Xuan Zhu, Jing-Ya Sun, Jun-Lan Lu, Jia-Kai Zeng, Feng-Qing Wang, Fei-Fei Du, Xi-He Yang, Shu-Ning Ge, Rui-Min Huang, Wei-Wei Jia, Chuan Li

## Abstract

The potential of Dahuang to eliminate lung pathogens was often highlighted in *Wenyi Lun*. This investigation aimed to identify potential antiviral compounds of herbal component Dahuang (*Rheum palmatum* rhizomes and roots) of LianhuaQingwen capsule, with respect to their systemic exposure and lung reachability. Circulating Dahuang compounds were identified in human volunteers receiving LianhuaQingwen. The reachability of these compounds to SARS-CoV-2 3CL^pro^ was assessed by *in vitro* transport, metabolism, immunohistochemistry, and 3CL^pro^-biochemical studies. LianhuaQingwen contained 55 Dahuang constituents (0.01–2.08 μmol/day), categorized into eight classes. Only three compounds rhein (**3**), methylisorhein (**10**; a new Dahuang anthraquinone), and 4-*O*-methylgallic acid (**M42_M2_**) exhibited significant systemic exposure in humans. Two intestinal absorption mechanisms for **3** and **10** were proposed: active intestinal uptake of **3**/**10** by human TAUT/ASBT and human MRP1/3/4, and intestinal lacate-phlorizin hrdrolyase-mediated hydrolysis of rhein-8-*O*-β-D-glucoside (**9**), followed by the transporter-mediated absorption of released **3**. Targeted reachability of circulating **3**/**10** could be achieved as rat orthologues of human ASBT/TAUT was observed in alveolar and bronchial epithelia. These compounds exhibited potential ability to inhibit the 3CL^pro^ enzyme responsible for coronaviral replication. Notably, Dahuang anthraquinones and tannins varied greatly in pharmacokinetics between humans and rats after dosing LianhuaQingwen. This investigation, along with such investigations of other components, has implications for precisely defining the therapeutic benefits of Dahuang-containing medicines.

## 1. Introduction

Dahuang (Rhubarb) has been used in traditional Chinese medicine for over 2000 years. Its medicinal properties and various applications were first recorded in the *Divine Husbandman’s Classic of Materia Medica* (*Shen Nong Ben Cao Jing*) during the Han Dynasty. The medicinal herb is obtained from the dried roots and rhizomes of three species: *Rheum palmatum* L., *Rheum tanguticum* Maxim. ex Balf., and *Rheum officinale* Baill, which are included in the 2020 Chinese Pharmacopeia [1, 2]. According to the *Epidemic Febrile Disease* (*Wenyi Lun*) in the Ming Dynasty, Dahuang was used to eliminate pestilential qi due to its penetrating properties without detainment [3]. This theory proposed that “epidemic disease” was caused by the infection of pestilential qi, which referred to an exogenous disease pathogen that was highly contagious and had an unpredictable epidemic pattern. Modern research provided evidence suggesting that Dahuang extracts possessed inhibitory activity against the 3-chymotrypsin-like protease (3CL^pro^) of severe acute respiratory syndrome coronavirus (SARS-CoV) [4]. Furthermore, an integrated computational study revealed that 13 natural anthraquinones, including chrysophanol, emodin, aloe-emodin, rhein, and alterporriol Q, exhibited promising potential as inhibitors of 3CL^pro^ of SARS-CoV-2 [5]. The application and findings indicated that Dahuang compounds might possess promising antiviral properties in humans. Additionally, the rapid development of modern science unraveled numerous pharmacological activities associated with Dahuang, including its purgative effects, anti-inflammatory properties, hepatoprotective benefits, and positive impacts on the gallbladder. Dahuang was used in clinical settings to treat constipation, severe acute pancreatitis, sepsis, and chronic liver and kidney diseases due to its extensive pharmacological effects [1, 6–8]. Dahuang has mainly been used for medicinal purposes in Asia, but it often refers to a few edible Dahuang in Europe and the Middle East [6, 7]. Concerns regarding the safety of Dahuang have arisen due to reports suggesting potential liver and kidney toxicity associated with long-term high-dose administration of Dahuang in rats, prompting global scrutiny on its safety profile [1, 6, 9].

LianhuaQingwen capsule, a traditional Chinese medicine that contains Dahuang, has been widely used in China for the treatment of acute respiratory viral infections caused by various viruses, including SARS-CoV, MERS-CoV, influenza A virus (including H1N1 and H7N9), and more recently, SARS-CoV-2 [10, 11]. Several clinical investigations reported the potential of the capsule as a valuable treatment option for both COVID-19 and influenza patients. Its ability to improve viral clearance, alleviate symptoms, and reduce inflammatory markers makes it a promising addition to conventional care [12–16]. In a recent prospective, multicenter, randomized controlled trial of 284 patients with COVID-19, adding LianhuaQingwen to conventional care further improved the recovery rate of symptoms, shortened the time to symptom recovery, and improved the recovery of chest radiologic abnormalities (*P* < 0.05 for all) [12]. Treatment of the capsule in influenza patients was found to reduce the levels of serum inflammatory factors C-reactive protein, IL-6, and procalcitonin [16]. In a cell-based study, LianhuaQingwen demonstrated inhibition of SARS-CoV-2 replication and induced abnormal particle morphology of the virion [17]. Additionally, several circulating constituents of LianhuaQingwen, including rhein, forsythoside A, forsythoside I, and neochlorogenic acid, were identified as inhibitors of the host target cell-surface protein known as angiotensin converting enzyme 2 receptor [18], to which the SARS-CoV-2 spike protein binds.

LianhuaQingwen capsule is derived from 13 herbal components and possesses complex chemical composition. For systemic treatment of acute respiratory viral infection with LianhuaQingwen, identification of circulating compounds in unchanged and/or metabolized forms is vital for facilitating their access to the loci responsible for therapeutic effects. To this end, comprehensive understanding of chemical composition of LianhuaQingwen and better evaluation of pharmacokinetics, disposition, and targeted reachability of its bioavailable compounds are vital for translating the potential benefits of the capsule into therapeutic application. Successful identification of such constituents necessitates a comprehensive approach to the multi-compound pharmacokinetic investigation of herbal medicine such that the potentially important herbal compounds are singled out with their accurate pharmacokinetic and disposition data and that no such compound is missed (in a few words, ‘precision without omission’) [19–30]. In our ongoing series of multi-compound pharmacokinetic inveatigations on LianhuaQingwen, we have previously conducted research on the constituents originating from the herbal component Gancao in humans after dosing the capsule [20]. In this current investigation, our focus is on the component Dahuang, with the aim of identifying the compounds that are likely to have therapeutic significance. Dahuang is extensively used as an herb in Chinese herbal medicines as well as dietary products. Therefore, further research is necessary to elucidate the specific compounds responsible for its pharmacological effects. This research has important implications for the safe utilization of Dahuang-containing herbal medicines. Specifically, this pharmacokinetic investigation aimed to identify bioavailable Dahuang compounds that have systemic exposure and targeted reachability, and are likely to influence the therapeutic outcomes of Dahuang-containing herbal medicines, such as LianhuaQingwen.

## 2. Materials and methods

A detailed description of materials and experimental procedures is provided in the Supplementary Materials and methods.

### 2.1. Study materials

LianhuaQingwen capsules are produced by Shijiazhuang Yiling Pharmaceutical Co., Ltd and have a Chinese NMPA drug ratification number of GuoYaoZhunZi-Z20040063. Samples of LianhuaQingwen capsules were obtained from the manufacturer, along with individual samples of the herbal components, and stored at -20°C until analysis to assess their chemical composition and lot-to-lot variability. Purified compounds found in *Rheum* species were obtained commercially (Table S1). One such compound, 3,8-dihydroxy-1-methylanthraquinone-2-carboxylic acid (methylisorhein), was extracted and isolated from Dahuang (*R. palmatum* rhizomes and roots) (Table S2).

Caco-2 cells and HEK-293 cells were obtained from American Type Culture Collection (Manassas, VA, USA). Various human transporter cDNAs and human enzyme cDNA lactase-phlorizin hydrolase (LPH) were constructed commercially. Inside-out membrane vesicles expressing human ATP-binding cassette transporter were purchased from GenoMembrane (Kanazawa, Japan). Pooled human liver microsomes (HLM) and pooled human liver cytosol (HLC) were obtained from Corning Gentest (Woburn, MA, USA). Commercially available positive substrates for these transporters and enzymes were also obtained.

### 2.2. Human pharmacokinetic study

A pharmacokinetic study on LianhuaQingwen capsule was conducted at Hebei Yiling Hospital (Shijiazhuang, Hebei Province, China). The study was approved by the hospital’s ethics committee and registered with the Chinese Clinical Trials Registry (ChiCTR1900021460), following the Declaration of Helsinki. Fourteen healthy volunteers participated in the study after providing written informed consent. The study details have been previously described [20]. Briefly, the participants received a single dose of 12 capsules of LianhuaQingwen on day 1 and days 3-8. Blood samples were collected before and at various time points after dosing on day 1, and urine samples were collected within specific time periods. On days 3-7, blood samples were collected before and 12 hours after dosing. On day 8, both blood and urine samples were collected according to the respective time schedules on day 1. The samples were treated and stored for analysis by liquid chromatography/mass spectrometry at −70°C.

### 2.3. Supportive in vitro transport and metabolism studies

#### 2.3.1. Transport study using Caco-2 cell monolayers

The membrane permeability of Dahuang compounds, including chrysophanol (**1**; 5 μmol/L), emodin-8-*O*-β-D-glucoside (**2**; 50 μmol/L), rhein (**3**; 5 μmol/L), chrysophanol-8-*O*-β-D-glucoside (**4**; 50 μmol/L), emodin (**5**; 5 μmol/L), physcion-8-*O*-β-D-glucoside (**6**; 50 μmol/L), chrysophanol-1-*O*-β-D-glucoside (**7**; 50 μmol/L), physcion (**8**; 5 μmol/L), rhein-8-*O*-β-D-glucoside (**9**; 50 μmol/L), methylisorhein (**10**; 5 μmol/L), and aloe-emodin (**11**; 5 μmol/L), was assessed using Caco-2 cell monolayers. The cells were cultured and the integrity of the cell monolayers was measured. The apparent permeability coefficient (*P*_app_) of each test compound was calculated (Supplementary Materials and methods). The test compounds were classified into three categories based on their *P*_app_ values: low, intermediate, or high membrane permeability [21]. The efflux ratio (EfR) was calculated and used to determine the possible involvement of a transporter-mediated efflux mechanism. Verapamil, MK571, and novobiocin were added to inhibit ABC transporters multidrug resistance protein 1 (MDR1), multidrug resistance-associated protein 2 (MRP2), and breast cancer resistance protein (BCRP). Atenolol and antipyrine were used as low and high permeability reference compounds, respectively, while rhodamine 123, sulfasalazine, and estrone-3-sulfate were used as probe substrates of MDR1, MRP2, and BCRP, respectively. The experimental details have been previously described [31].

#### 2.3.2. Transport studies using cells transfected with SLC transporters or membrane vesicles expressing ABC transporters

Cellular uptake of chrysophanol (**1**; 5 μmol/L), emodin-8-*O*-β-D-glucoside (**2**; 50 μmol/L), rhein (**3**; 5 μmol/L), chrysophanol-8-*O*-β-D-glucoside (**4**; 50 μmol/L), emodin (**5**; 5 μmol/L), physcion-8-*O*-β-D-glucoside (**6**; 50 μmol/L), chrysophanol-1-*O*-β-D-glucoside (**7**; 50 μmol/L), physcion (**8**; 5 μmol/L), rhein-8-*O*-β-D-glucoside (**9**; 50 μmol/L), methylisorhein (**10**; 5 μmol/L), aloe-emodin (**11**; 5 μmol/L), gallic acid (**42**; 5 μmol/L), and 4-*O*-methylgallic acid (**M42_M2_**; 5 μmol/L) was assessed using human apical sodium-dependent bile acid transporter (ASBT)-, taurine transporter (TAUT)-, organic anion-transporting polypeptide 2B1 (OATP2B1)-, organic anion transporter 1 (OAT1)-, OAT2-, OAT3-, OAT4-, organic cation transporter 2 (OCT2)-, peptide transporter 1 (PEPT1)-, and PEPT2-transfected HEK-293 cells. In addition, vesicular transport of **3** (5 μmol/L) and **10** (5 μmol/L) was assessed using vesicles expressing human MDR1, BCRP, MRP1, MRP2, MRP3, or MRP4. All the incubation samples were analyzed by liquid chromatography/mass spectrometry. The experimental details have been previously described [32, 33].

#### 2.3.3. Metabolism studies

The deglycosylation of emodin-8-*O*-β-D-glucoside (**2**), chrysophanol-8-*O*-β-D-glucoside (**4**), physcion-8-*O*-β-D-glucoside (**6**), chrysophanol-1-*O*-β-D-glucoside (**7**), or rhein-8-*O*-β-D-glucoside (**9**), each at 1 μmol/L, was assessed using recombinant human LPH. Glucuronidation, sulfation, and oxidation of rhein (**3**) or methylisorhein (**10**), each at 1 μmol/L, were assessed using UDPGA-fortified HLM, PAPS-fortified HLC, and NADPH-fortified HLM. The detailed incubation conditions were described previously [30].

### 2.4. Supportive rat pharmacokinetic studies

Rat pharmacokinetic study on LianhuaQingwen capsule was conducted at Laboratory Animal Center of Shanghai Institute of Materia Medica (Shanghai, China). Three studies were approved by the Institutional Animal Care and Use Committee, following the Guidelines for Ethical Treatment of Laboratory Animals by the Ministry of Science and Technology of China. Femoral artery cannulation or bile duct cannulation was performed to collect rat blood or bile, respectively [23]. In these three rat studies, all the rats were given an oral dose of LianhuaQingwen at 3.78 g/kg. The first study involved the collection of serial blood samples before and at various time points after dosing. The blood samples were treated and centrifuged to produce plasma samples. The second study involved the collection of serial urine samples within specific time periods after dosing. The third study involved the collection of serial bile samples within specific time periods after dosing. The samples were stored for analysis by liquid chromatography/mass spectrometry at −70°C.

### 2.5. Assay for composition analysis of Dahuang constituents in LianhuaQingwen

The composition analysis of LianhuaQingwen for constituents originating from the component Dahuang was conducted using liquid chromatography/mass spectrometry. A literature-derived list of candidate compounds was used to guide the analysis, and a comprehensive detection approach was employed to identify all constituents present in the sample. The detected constituents underwent characterization and quantification for ranking and grading. Fourteen capsule lots were analyzed to assess inter-lot variability.

### 2.6. Bioanalytical assays for analyses of Dahuang compounds and other study compounds

The pharmacokinetics of Dahuang compounds in LianhuaQingwen administration was investigated through two types of bioanalyses. The first type involved profiling unchanged and metabolized Dahuang compounds in human/rat samples. This was accomplished utilizing the Acquity ultra performance liquid chromatographic separation module/Waters Synapt G2 mass spectrometer to detect, characterize, and quantify the compounds. The second type involved quantifying specific Dahuang compounds in human/rat samples and *in vitro* samples using an Applied Biosystems Sciex Triple Quad 5500 mass spectrometer interfaced with an Agilent 1290 Infinity Ⅱ LC system. The concentrations of various reference compounds in *in vitro* study samples were analyzed using liquid chromatography/mass spectrometry. Assay validation, implemented according to the European Medicines Agency Guideline on bioanalytical method validation (2012), demonstrated that the quantification assays developed were reliable and reproducible for the intended use, despite no internal standard being used.

### 2.7. In vitro assessment of SARS-CoV-2 3CL^pro^ protease inhibition

The proteolytic activity of the 3-chymotrypsin-like protease (3CL^pro^) of SARS-CoV-2 was assessed with a fluorescence resonance energy transfer (FRET) assay. The activity of recombinant SARS-CoV-2 3CL^pro^ was evaluated in cleaving a synthetic fluorogenic peptide substrate MCA-AVLQSGFR-Lys(Dnp)-Lys-NH_2_, which was derived from the N-terminal auto-cleavage sequence from the viral protease. The cleaved MCA peptide fluorescence was measured by a SpectraMax M2 microplate reader (Molecular Devices; CA, USA) at an excitation wavelength of 325 nm and an emission wavelength of 393 nm. The assay reaction buffer contained 50 mM Tris-HCl (pH 7.3), 1 mM EDTA, and 20 μM peptide substrate. Enzymatic reactions were initiated with the addition of 100 nM SARS-CoV-2 3CL^pro^ and allowed to proceed at 37 °C for 10 min (under linear cleavage condition). The Michaelis-Menten constant of the peptide substrate for SARS-CoV-2 3CL^pro^ was measured to determine the substrate concentration used in the following inhibition assessment. Ebselen, a positive inhibitor of SARS-CoV-2 3CL^pro^, decreased the fluorescence. The inhibition potencies of the test Dahuang compounds, such as rhein (**3**), methylisorhein (**10**), aloe-emodin (**11**), 4-*O*-methylgallic acid (**M42_M2_**), and gallic acid (**42**), were initially screened at a concentration of 100 μmol/L. The Dahuang compounds that showed ≥ 50% inhibition in the initial screening were further evaluated at multiple concentrations for their half-maximal inhibitory concentrations (IC_50_) for SARS-CoV-2 3CL^pro^. For the IC_50_ assessment, the recombinant protease was incubated with the peptide substrate at 1.4 μmol/L (the *K*_m_ for SARS-CoV-2 3CL^pro^) in the presence or absence of the test Dahuang compound.

### 2.8. Western blot

HEK-293 cells were transfected with empty vector, human Flag-ASTB, human Flag-TAUT, rat Flag-ASBT, or rat Flag-TAUT plasmids for 48 hours. After that, the cells were harvested and lysed with RIPA lysis buffer (pH 7.5) containing 150 mM NaCl, 1 mM Na_3_VO_4_, 50 mM Tris-HCl, 25 mM NaF, 1% NP-40, 0.5% sodium deoxycholate, 0.1% SDS, and 1% phosphatase inhibitor cocktails. Rat lung, bronchus, liver, and intestine were collected and lysed in the same RIPA lysis buffer using a grinder. Subsequently, 20 μg of the aforementioned proteins were loaded onto SDS-PAGE and transferred to nitrocellulose membranes (Millipore, Billerica, MA, USA). The membranes were then incubated overnight at 4°C with primary antibodies against ASBT (1:1000, 25245-1-AP; Proteintech, Wuhan, Hubei Province, China) or TAUT (1:1000, PA5-98161; Invitrogen, Waltham, MA, USA). For detection, the membranes were treated with secondary antibodies conjugated to horseradish peroxidase (Jackson, West Grove, PA, USA) and the immunoreactive proteins were visualized using the West Pico PLUS Chemiluminescent Substrate (Thermo, Waltham, MA, USA) and imaged using chemiluminescent imaging.

### 2.9. Immunohistochemistry

Rat lung and intestine tissues were fixed in 4% paraformaldehyde and embedded in paraffin. The tissues were then sectioned into 5 µm thick paraffin sections and deparaffinized. Antigen retrieval was performed using Tris-EDTA (pH 9.0). For immunohistochemistry staining, the sections were incubated with primary antibodies against ASBT (1:100, AB203205; Abcam, Cambridge, UK) or TAUT (1:500, PA5-98161; Invitrogen, Waltham, MA, USA), followed by incubation with biotinylated goat anti-rabbit IgG as secondary antibodies. Signals were amplified using a DAB kit (Maixin Bio, Fuzhou, Fujian Province, China) and counterstained with hematoxylin.

### 2.10. Data Processing

After conducting the composition analysis, the detected and characterized Dahuang constituents were ranked and graded into different levels based on their daily doses. The daily compound dose was calculated by multiplying the compound level in LianhuaQingwen and the capsule’s label daily dose of 4.2 g/day. Noncompartmental analysis was used to estimate pharmacokinetic parameters of Dahuang compounds. Renal clearance ratio (*R*_rc_) was calculated using renal clearance, glomerular filtration rate [34], and unbound fraction in human plasma. Data are expressed as mean ± standard deviation, and statistical analysis was performed using a two-tailed unpaired Student t-test. A value of *P* < 0.05 was considered statistically significant.

## 3. Results

### 3.1. Constituents originating from Dahuang and their abundance in LianhuaQingwen

Based on the composition analysis conducted, a total of 55 constituents that originated from the Dahuang component were detected and characterized in samples of LianhuaQingwen (Fig. 1 and Table S3). These constituents were divided into eight classes, including anthraquinones (**1**–**22**), anthrones (**31**–**34**), tannins (**41**–**52**), flavonoids (**61**–**64**), naphthalenes (**71**–**73**), stilbenes (**81**–**84**), pyranones (**91**–**94**), and phenylbutanones (**101** and **102**). All of these constituents had a compound dose of ≥ 0.01 μmol/day. Notably, these constituents were not detected in any other components of LianhuaQingwen, except for gallic acid (**42**) which was also detected in the component Hongjingtian (*R. crenulata* rhizomes and roots). The major constituents of Dahuang were identified as chrysophanol (**1**), emodin-8-*O*-β-D-glucoside (**2**), rhein (**3**), chrysophanol-8-*O*-β-D-glucoside (**4**), emodin (**5**), and physcion-8-*O*-β-D-glucoside (**6**). These major constituents had a compound dose of 1.0–2.2 µmol/day. Moreover, LianhuaQingwen also exhibited lot-to-lot variations of 8.3%–12.1% for the major anthraquinones (**1**–**6**) and 6.6%–55.3% for the rest of the minor constituents.

**Fig. 1.**
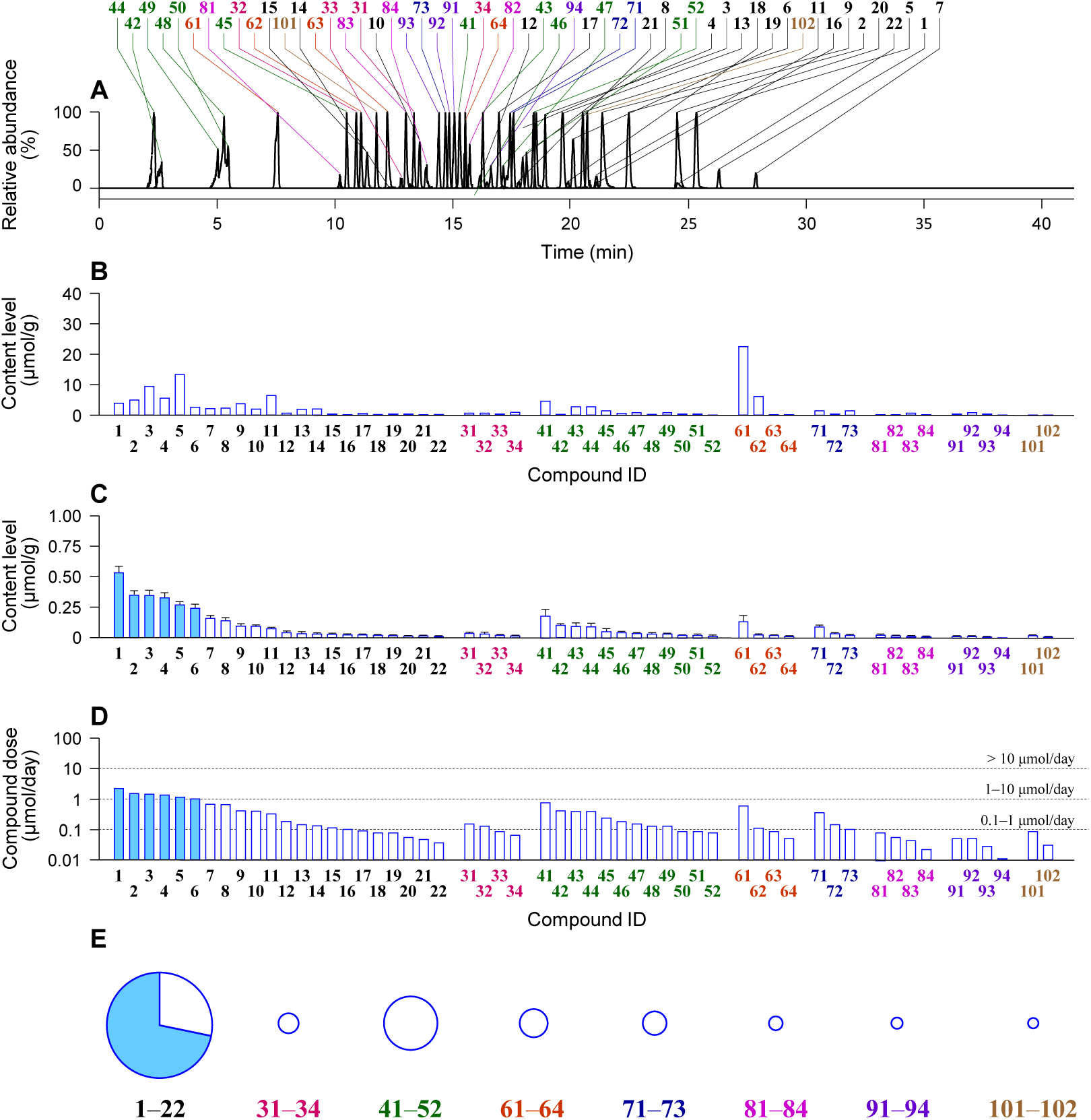
LianhuaQingwen constituents originating from Dahuang. (A) stacked liquid chromatograms of anthraquinones (**1**–**22**), anthrones (**31**–**34**), tannins (**41**–**52**), flavonoids (**61**–**64**), naphthalenes (**71**–**73**), stilbenes (**81**–**84**), pyranones (**91**–**94**), and phenylbutanones (**101** and **102**), detected by mass spectrometry, in a typical sample of LianhuaQingwen; (B) constituents detected in a sample of raw material of Dahuang (*Rheum palmatum* rhizomes and roots); (C) mean content levels of the constituents detected in samples of 14 lots of LianhuaQingwen; (D) daily doses of the constituents from the lot A1802055 of LianhuaQingwen at the label daily dose 4.2 g/day; (E) percentage daily doses of anthraquinones, anthrones, tannins, flavonoids, naphthalenes, stilbenes, pyranones, and phenylbutanones in their respective total daily dose of all the constituents of the class detected in LianhuaQingwen. **1**, chrysophanol; **2**, emodin-8-*O*-β-D-glucoside; **3**, rhein; **4**, chrysophanol-8-*O*-β-D-glucoside; **5**, emodin; **6**, physcion-8-*O*-β-D-glucoside; and **10**, methylisorhein. The names of the other 48 constituents originating from Dahuang are shown in Table S3. The details of composition analysis of LianhuaQingwen for constituents originating from Dahuang are described in the Supplementary Materials and methods.

### 3.2. Major circulating Dahuang compounds after dosing LianhuaQingwen and their plasma pharmacokinetics: interspecies differences between humans and rats

Only two classes of compounds, i.e., anthraquinones and tannins, had significant systemic exposure in humans following administration of LianhuaQingwen (Fig. 2 and Table 1), despite the presence of 55 Dahuang constituents of eight classes in the capsule. Human systemic exposure to anthraquinones varied greatly, with unchanged rhein (**3**) and methylisorhein (**10**), rather than their metabolites, being the most abundant in circulation. The other anthraquinones, including chrysophanol (**1**), emodin-8-*O*-β-D-glucoside (**2**), chrysophanol-8-*O*-β-D-glucoside (**4**), emodin (**5**), physcion-8-*O*-β-D-glucoside (**6**), chrysophanol-1-*O*-β-D-glucoside (**7**), physcion (**8**), rhein-8-*O*-β-D-glucoside (**9**), and aloe-emodin (**11**), were either not detectable or present in negligible levels in human plasma, either as unchanged compounds or metabolites. The major circulating compound of tannins was 4-*O*-methylgallic acid (**M42_M2_**), which was a methylated metabolite of gallic acid (**42**), while **42** and its other methylated metabolite 3-*O*-methylgallic acid (**M42_M1_**) were detected at low levels in plasma (Fig. 2). All circulating compounds were also detected in human urine.

**Fig. 2.**
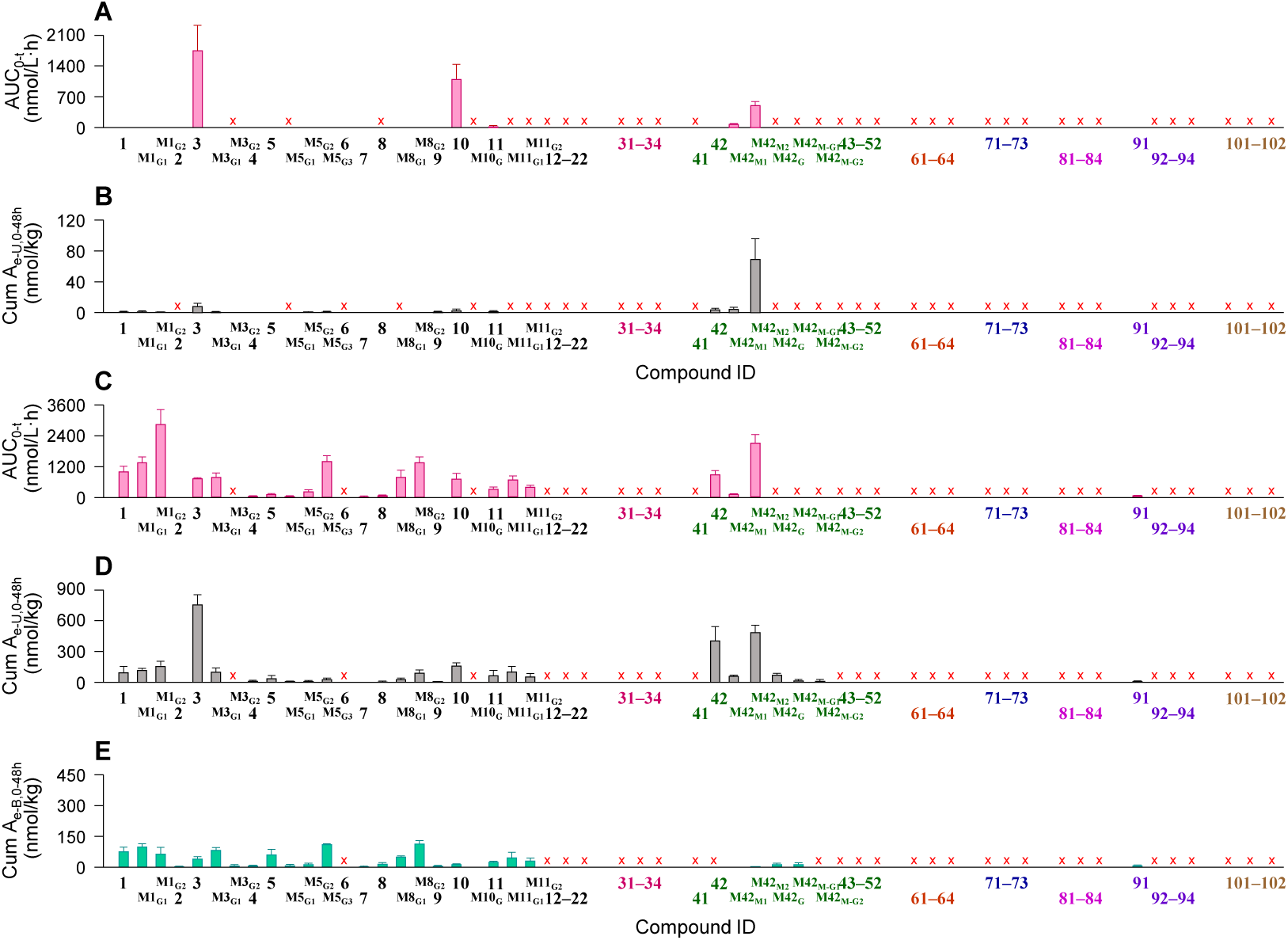
Dahuang compounds detected in healthy volunteers and rats after orally dosing LianhuaQingwen. (A and C) systemic exposure data; (B and D) renal excretion data; (E) hepatobiliary excretion data. The dose for human volunteers was 12 capsules/person (A and B), while that for rats was 3.78 g/kg (C–E). Both unchanged and metabolized Dahuang compounds were detected. **3**, rhein; **10**, methylisorhein, **M42M2**, 4-*O*-methylgallic acid. The names of the other Dahuang compounds detected are shown in Table 1.

**Table 1.** Dahuang compounds, unchanged and metabolized, detected in human samples after orally dosing LianhuaQingwen.

| Compound (ID) | LC/TOF-MS<sup>E</sup> data |  |  |  | Molecular formula | Presence in human samples |
| --- | --- | --- | --- | --- | --- | --- |
|  | *t*<sub>R</sub> (min) | [M–H]<sup>–</sup> (*m/z*) | Mass error (ppm) | Fragmentation profile (*m/z*) |  |  |
| Chrysophanol (**1**) | 27.14 | 253.0511 | 4.0 | 225.0569, 183.0091 | C<sub>15</sub>H<sub>10</sub>O<sub>4</sub> | Plasma, urine |
| Chrysophanol-1-*O*-glucuronide (**M1<sub>G1</sub>**) | 19.24 | 429.0815 | –1.6 | 253.0507, 225.0492, 183.0077 | C<sub>21</sub>H<sub>18</sub>O<sub>10</sub> | Plasma, urine |
| Chrysophanol-8-*O*-glucuronide (**M1<sub>G2</sub>**) | 19.41 | 429.0822 | 0 | 253.0505, 225.0708, 183.0178 | C<sub>21</sub>H<sub>18</sub>O<sub>10</sub> | Plasma, urine |
| Emodin-*O*-β-D-glucoside (**2**) | 18.94 | 431.0966 | –2.8 | 269.0443, 225.1192 | C<sub>21</sub>H<sub>20</sub>O<sub>10</sub> | Plasma |
| Rhein (**3**) | 23.61 | 283.0247 | 1.4 | 239.0348, 211.0422, 183.0411 | C<sub>15</sub>H<sub>18</sub>O<sub>6</sub> | Plasma, urine |
| Rhein-*O*-glucuronide-1 (**M3<sub>G1</sub>**) | 15.23 | 459.0563 | –0.2 | 283.0244, 239.0339 | C<sub>21</sub>H<sub>16</sub>O<sub>12</sub> | Plasma, urine |
| Rhein-*O*-glucuronide-2 (**M3<sub>G2</sub>**) | 16.93 | 459.0555 | –2.0 | 283.0234, 239.0352 | C<sub>21</sub>H<sub>16</sub>O<sub>12</sub> | Urine |
| Emodin (**5**) | 26.01 | 269.0455 | 1.9 | 241.0667, 225.0515, 197.0307 | C<sub>15</sub>H<sub>10</sub>O<sub>5</sub> | Plasma, urine |
| Emodin-*O*-glucuronide (**M5<sub>G2</sub>**) | 19.53 | 445.0764 | –1.6 | 269.0394, 225.0772 | C<sub>21</sub>H<sub>18</sub>O<sub>11</sub> | Plasma, urine |
| Emodin-3-*O*-glucuronide (**M5<sub>G3</sub>**) | 21.24 | 445.0776 | 1.1 | 269.0451, 225.0622 | C<sub>21</sub>H<sub>18</sub>O<sub>11</sub> | Plasma, urine |
| Physcion (**8**) | 28.60 | 283.0605 | –0.4 | 240.0323 | C<sub>16</sub>H<sub>12</sub>O<sub>5</sub> | Urine |
| Physcion-1-*O*-glucuronide (**M8<sub>G1</sub>**) | 20.92 | 459.0924 | –0.7 | 283.0599 | C<sub>22</sub>H<sub>20</sub>O<sub>11</sub> | Plasma |
| Physcion-8-*O*-glucuronide (**M8<sub>G2</sub>**) | 21.42 | 459.0923 | –0.9 | 283.0607 | C<sub>22</sub>H<sub>20</sub>O<sub>11</sub> | Plasma, urine |
| Methylisorhein (**10**) | 21.93 | 297.0392 | –2.4 | 253.0485 | C<sub>16</sub>H<sub>10</sub>O<sub>6</sub> | Plasma, urine |
| Aloe-emodin (**11**) | 21.81 | 269.0457 | 2.6 | 240.0364, 211.1301, 183.0977 | C<sub>15</sub>H<sub>10</sub>O<sub>5</sub> | Plasma, urine |
| Gallic acid (**42**) | 3.73 | 169.0137 | 0 | 125.0227, 78.9810 | C<sub>7</sub>H<sub>6</sub>O<sub>5</sub> | Plasma, urine |
| 3-*O*-Methylgallic acid (**M42<sub>M1</sub>**) | 7.65 | 183.0297 | 2.2 | 168.0056, 139.0397, 124.0153 | C<sub>8</sub>H<sub>8</sub>O<sub>5</sub> | Plasma, urine |
| 4-*O*-Methylgallic acid (**M42<sub>M2</sub>**) | 8.05 | 183.0299 | 3.3 | 168.0064, 139.0371, 124.0163 | C<sub>8</sub>H<sub>8</sub>O<sub>5</sub> | Plasma, urine |
| 2,5-Dimethyl-7-hydroxychrome (**91**) | 16.38 | 189.0556 | 2.1 | 174.0307, 146.0338 | C<sub>11</sub>H<sub>10</sub>O<sub>3</sub> | Plasma, urine |

The details of human and rat studies and bioanalytical assay are described in the Supplementary Materials and methods.

The data presented in Fig. 2 highlights significant differences in the exposure profile of Dahuang anthraquinones between rats and humans when they received LianhuaQingwen. Although rhein (**3**) and methylisorhein (**10**) were the major circulating anthraquinones for both rats and humans, rats had additional circulating anthraquinones and their metabolites that either went undetected or were present in limited quantities in human plasma. These newly detected anthraquinones in rats included chrysophanol (**1**), chrysophanol-1-*O*-glucuronide (**M1_G1_**), chrysophanol-8-*O*-glucuronide (**M1_G2_**), rhein-*O*-glucuronide (**M3_G1_**), emodin-3-*O*-glucuronide (**M5_G3_**), physcion-1-*O*-glucuronide (**M8_G1_**), and physcion-8-*O*-glucuronide (**M8_G2_**). The major circulating compound in humans for tannins was 4-*O*-methylgallic acid (**M42_M2_**), while both **M42_M2_** and gallic acid (**42**) were the major circulating compounds in rats. These findings suggest that the systemic exposure to anthraquinones and tannins could differ considerably between rats and humans, and that such differences should be taken into account when evaluating the safety and efficacy of the Dahuang-containing herbal medicine in preclinical studies.

Table 2 summarizes the plasma pharmacokinetics of major circulating Dahuang compounds [rhein (**3**), methylisorhein (**10**), and 4-*O*-methylgallic acid (**M42_M2_**)] in humans who were given 12 capsules/day of LianhuaQingwen. As illustrated in Fig. 3, both **3** and **10** showed unimodal plasma concentration-time profiles, reaching their peak concentrations (*T*_peak_) between 1 and 3 h. Females had a higher *C*_max_ and AUC_0-24h_ for both **3** and **10** compared to males (*P* < 0.05), with all the data adjusted for body weight. There was no significant accumulation of **3** and **10** after administering LianhuaQingwen for seven consecutive days (P > 0.05). As shown in Table 2, renal clearance ratio (*R*_rc_) of **3** and **10** suggested that the renal excretion involved tubular secretion, which was mediated by the renal basolateral SLC uptake transporters OAT1, OAT2, and OAT3 (Tables 3 and S4). In addition, **10**, but not **3**, was also a substrate of the renal apical SLC uptake transporter OAT4, which could induce tubular reabsorption of **10**. The plasma concentration-time profiles of **M42_M2_** were similar to those of **3** and **10**, as demonstrated in Table 2 and Fig. 3. The *R*_rc_ of **M42_M2_** in Table 2 showed that its renal excretion involved tubular secretion, which was also mediated by OAT1, OAT2, and OAT3 (Table 3).

**Fig. 3.**
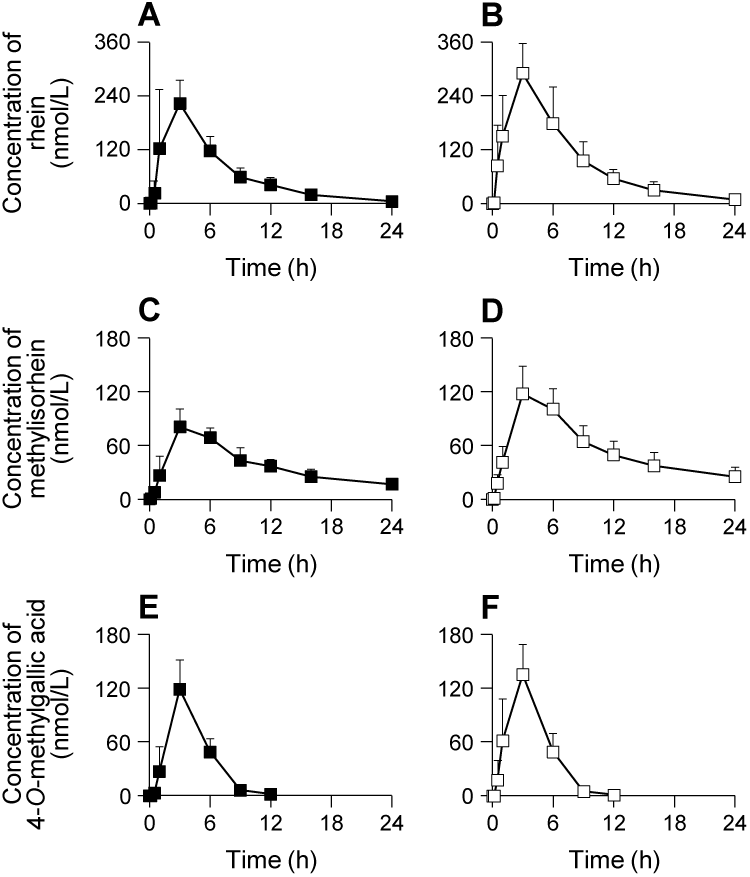
Plasma concentration-time profiles of Dahuang compounds in volunteers after dosing LianhuaQingwen at 12 capsules/person. (A and B), rhein; (C and D), methylisorhein; (E and F), 4-*O*-methylgallic acid. Solid dots, data of males; Open dots, data of females. Pharmacokinetic data of these compounds are shown Table 2.

**Table 2.**
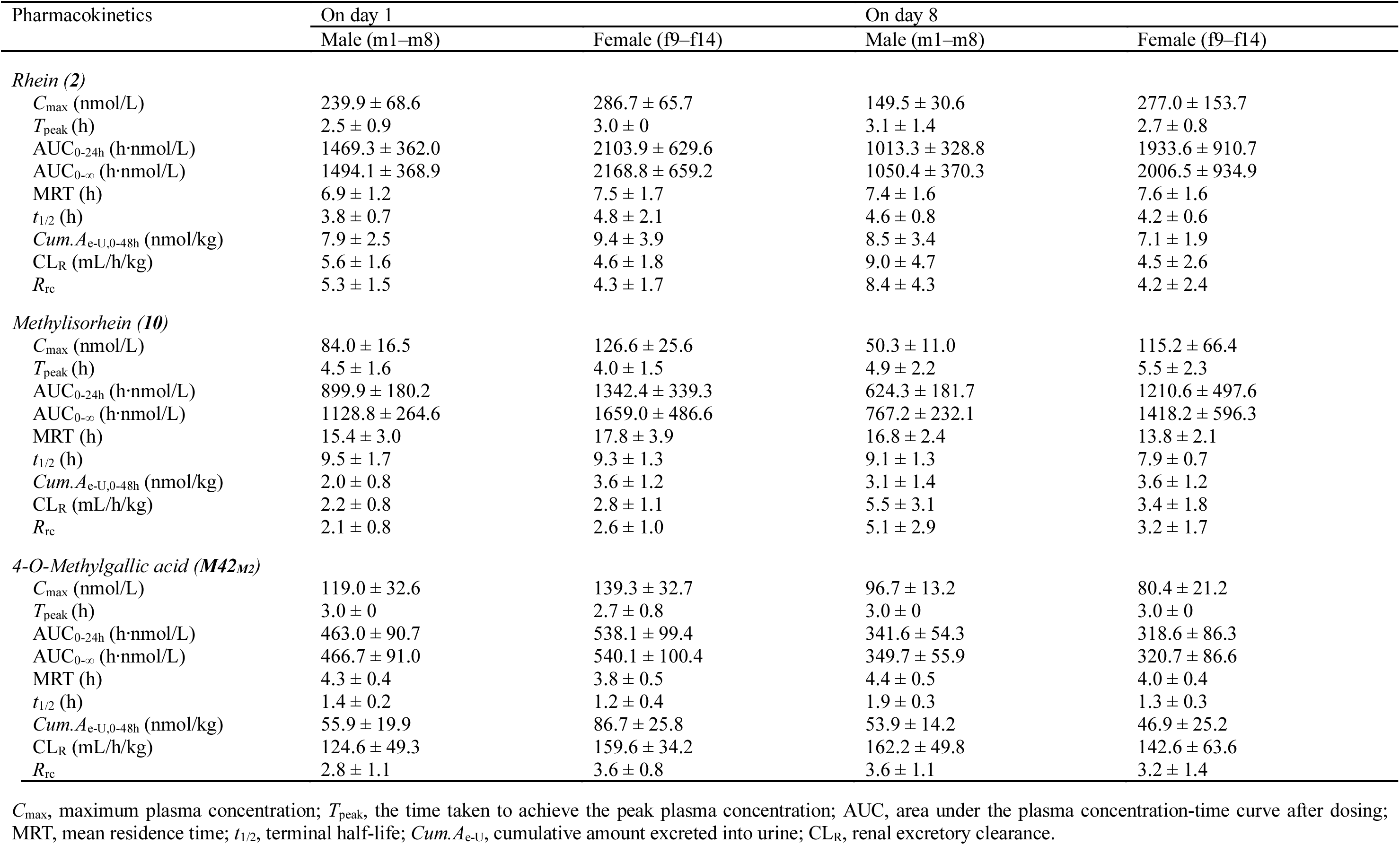
Pharmacokinetics of rhein (3), methylisorhein (10), and 4-*O*-methylgallic acid (M42M2) in volunteers orally receiving LianhuaQingwen.

**Table 3.** Passive diffusion of Dahuang compounds in mock HEK-293 cells and their net transport ratios in HEK-293 cells transfected with various human intestinal or renal transporters.

| Compound (ID) | Transport in mock cell<br>($\mu\text{L}/\text{min}/\text{mg}$ protein) | Net transport ratio | | | | | | | | | |
| --- | --- | --- | --- | --- | --- | --- | --- | --- | --- | --- | --- |
|  |  | TAUT | ASBT | OATP2B1 | PEPT1 | OAT1 | OAT2 | OAT3 | OAT4 | OCT2 | PEPT2 |
| Positive substrate | — | $7.2 \pm 1.5$ | $200 \pm 38$ | $12.3 \pm 2.4$ | $17.6 \pm 2.1$ | $476 \pm 57$ | $24.1 \pm 3.6$ | $4.5 \pm 0.3$ | $7.6 \pm 1.3$ | $14.5 \pm 2.6$ | $54.9 \pm 3.9$ |
| Chrysophanol (**1**) | $8.8 \pm 1.1$ | $1.0 \pm 0.1$ | $0.9 \pm 0.5$ | $0.9 \pm 0.3$ | $2.2 \pm 0.2$ | — | — | — | — | — | — |
| Emodin-8-*O*-β-D-glucoside (**2**) | $0.1 \pm 0.0$ | $2.1 \pm 0.3$ | $0.8 \pm 0.1$ | $3.6 \pm 0.1$ | $0.9 \pm 0.1$ | — | — | — | — | — | — |
| Rhein (**3**) | $0.2 \pm 0.0$ | $18.4 \pm 2.7$ | $5.9 \pm 0.7$ | $1.0 \pm 0$ | $0.7 \pm 0.1$ | $9.9 \pm 1.0$ | $8.4 \pm 0.3$ | $1.1 \pm 0.1$ | $1.8 \pm 0.3$ | $1.0 \pm 0.2$ | $1.7 \pm 0.3$ |
| Chrysophanol-8-*O*-β-D-glucoside (**4**) | $1.5 \pm 0.2$ | $0.8 \pm 0.1$ | $1.4 \pm 0.1$ | $2.0 \pm 0.4$ | $0.8 \pm 0.1$ | — | — | — | — | — | — |
| Emodin (**5**) | $27.8 \pm 1.6$ | $1.0 \pm 0.1$ | $1.7 \pm 0.4$ | $1.6 \pm 0.1$ | $0.8 \pm 0.2$ | — | — | — | — | — | — |
| Physcion-8-*O*-β-D-glucoside (**6**) | $6.0 \pm 1.2$ | $0.7 \pm 0.1$ | $1.0 \pm 0.3$ | $1.2 \pm 0.2$ | $0.8 \pm 0.1$ | — | — | — | — | — | — |
| Chrysophanol-1-*O*-β-D-glucoside (**7**) | $1.4 \pm 0.1$ | $1.3 \pm 0.4$ | $1.2 \pm 0.1$ | $0.7 \pm 0$ | $1.5 \pm 0.6$ | — | — | — | — | — | — |
| Physcion (**8**) | $9.4 \pm 2.4$ | $0.8 \pm 0.1$ | $1.8 \pm 0.4$ | $1.1 \pm 0.1$ | $1.2 \pm 0.2$ | — | — | — | — | — | — |
| Rhein-8-*O*-β-D-glucoside (**9**) | $0.02 \pm 0$ | $0.8 \pm 0.1$ | $0.7 \pm 0.4$ | $0.7 \pm 0$ | $0.9 \pm 0.1$ | — | — | — | — | — | — |
| Methylisorhein (**10**) | $0.1 \pm 0.0$ | $7.7 \pm 0.4$ | $3.1 \pm 0.3$ | $2.4 \pm 0.1$ | $2.2 \pm 0.1$ | $8.1 \pm 0.6$ | $11.8 \pm 0.3$ | $5.7 \pm 0.0$ | $4.9 \pm 1.1$ | $1.3 \pm 0.1$ | $1.2 \pm 0.3$ |
| Aloe-emodin (**11**) | $24.8 \pm 3.1$ | $1.1 \pm 0.3$ | $1.2 \pm 0.2$ | $1.0 \pm 0.1$ | $1.4 \pm 0.2$ | — | — | — | — | — | — |
| Gallic acid (**42**) | $0.01 \pm 0$ | $1.1 \pm 0.2$ | $1.2 \pm 0.4$ | $1.6 \pm 0.5$ | $0.5 \pm 0$ | — | — | — | — | — | — |
| 4-*O*-methylgallic acid (**M42M2**) | $0.03 \pm 0$ | — | — | — | $0.9 \pm 0.3$ | $73.2 \pm 3.3$ | $20.1 \pm 0.8$ | $5.5 \pm 0.6$ | $1.5 \pm 0.1$ | $0.6 \pm 0.2$ | $1.1 \pm 0.2$ |

For assessment of cellular uptake mediated by intestinal or renal transporters, differential transport rates between transfected cells and mock cells were defined as a net transport ratio, which represents the mean ± standard deviation ( $n = 3$ ). A net transport ratio $> 3$ suggested a positive result. Positive substrates were taurocholic acid (ASBT), [ $\text{H}^3$ ]-taurine (TAUT), estrone-3-sulfate (OATP2B1/OAT3/OAT4), glycylsarcosine (PEPT1/PEPT2), *para*-aminohippuric acid (OAT1), prostaglandin $\text{F}_{2a}$ (OAT2), tetraethylammonium (OCT2) and their final concentrations were $20 \mu\text{mol}/\text{L}$ with incubation time of 10 min.

### 3.3 Factors governing the differential human systemic exposure to Dahuang anthraquinones: transporter-mediated intestinal absorption and LPH-mediated pre-absorption metabolism

Membrane permeability was assessed using Caco-2 cell monolayer to understand the factors governing differential human systemic exposure to the anthraquinones. Rhein (**3**) and methylisorhein (**10**) showed moderate membrane permeability (Table 4). Estimated water solubility of these two compounds (Table S5), in relation to their respective compound doses from LianhuaQingwen and Caco-2-based permeability, was insufficient to achieve adequate intestinal absorption [35, 36]. Intestinal penetration of **3** and **10** could also be limited by human intestinal apical MDR1-, BCRP-, and MRP2-mediated efflux into the intestinal lumen (Table 4 and Table S6). However, human intestinal apical ASBT and TAUT could enhance intestinal absorption of these compounds into intestinal epithelia (Table 3), and human intestinal basolateral MRP1, MRP3, and MRP4 could mediate the compounds’ efflux into blood (Table S6). Rhein-8-*O*-β-D-glucoside (**9**), which had poor membrane permeability for intestinal absorption, was not a substrate of ASBT or TAUT. However, this anthraquinone glucoside was found to be a substrate of human intestinal LPH, which could facilitate the pre-absorption deglycosylation of **9** into the absorbable **3**. *In vitro* metabolism studies suggested that **3** and **10** were resistant to human hepatic glucuronidation, sulfation, or oxidation (Fig. S1).

**Fig. 4.**
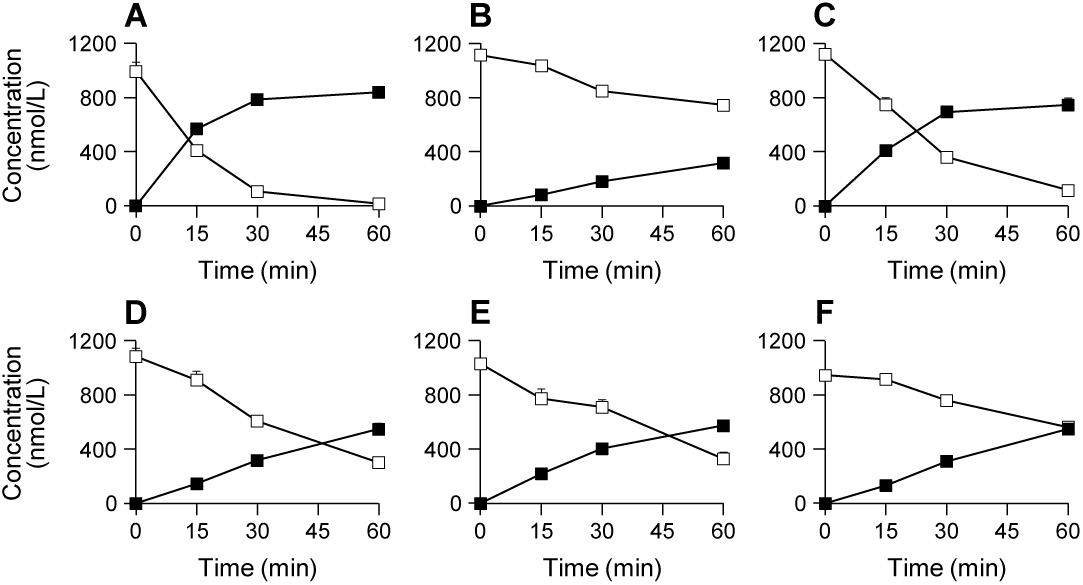
*In vitro* human LPH-mediated deglycosylation of Dahuang anthraquinone glycosides. Deglycosylation of the positive substrate phlorizin to phloretin (A) and Dahuang compounds emodin-8-*O*-β-D-glucoside to emodin (B), chrysophanol-8-*O*-β-D-glucoside to chrysophanol (C), physcion-8-*O*-β-D-glucoside to physcion (D), chrysophanol-1-*O*-β-D-glucoside to chrysophanol (E), and rhein-8-*O*-β-D-glucoside to rhein (F). Solid dots, anthraquinone glycosides; open dots, the respective free anthraquinones. Other *in vitro* metabolism data of Dahuang compounds are shown in Supplementary Fig. S1.

**Table 4.** Apparent permeability for the investigational Dahuang compounds in Caco-2 cell monolayers.

| Compound | Without inhibitors |  |  | With inhibitors |  |  |
| --- | --- | --- | --- | --- | --- | --- |
| | $P_{app}$ ($\times 10^{-6}$ cm/s) | | EfR | $P_{app}$ ($\times 10^{-6}$ cm/s) | | EfR |
|  | Apical→Basolateral | Basolateral→Apical |  | Apical→Basolateral | Basolateral→Apical |  |
| Atenolol | 0.8 ± 0.1 | 0.7 ± 0.1 | 0.9 | — | — | — |
| Antipyrine | 33.1 ± 7.8 | 36.6 ± 6.4 | 1.1 | — | — | — |
| Rhodamine 123 | 0.8 ± 0.1 | 10.9 ± 1.2 | 14.0 | 0.3 ± 0.0 | 0.6 ± 0.0 | 1.9 |
| Sulfasalazine | 0.6 ± 0.0 | 10.7 ± 1.7 | 17.5 | 0.8 ± 0.0 | 1.6 ± 0.1 | 1.9 |
| Estrone-3-sulfate | 0.8 ± 0.2 | 7.4 ± 0.1 | 9.3 | 1.7 ± 0.1 | 1.4 ± 0.1 | 0.8 |
| Chrysophanol (1) | 2.0 ± 0.5 | 3.9 ± 0.1 | 2.0 | — | — | — |
| Emodin-8-*O*-β-D-glucoside (2) | 0.2 ± 0.0 | 2.7 ± 0.2 | 14.8 | 0.3 ± 0.1 | 0.9 ± 0.0 | 2.8 |
| Rhein (3) | 5.5 ± 0.6 | 34.2 ± 1.8 | 6.3 | 7.8 ± 0.9 | 17.0 ± 4.7 | 2.2 |
| Chrysophanol-8-*O*-β-D-glucoside (4) | 6.1 ± 0.2 | 25.6 ± 7.7 | 4.2 | 11.7 ± 0.9 | 10.2 ± 0.5 | 0.9 |
| Emodin (5) | 18.6 ± 1.3 | 16.5 ± 4.0 | 0.9 | — | — | — |
| Physcion-8-*O*-β-D-glucoside (6) | 0.6 ± 0.0 | 8.9 ± 1.3 | 14.9 | 1.2 ± 0.2 | 4.3 ± 0.9 | 3.5 |
| Chrysophanol-1-*O*-β-D-glucoside (7) | 3.8 ± 0.5 | 22.0 ± 2.0 | 5.8 | 9.0 ± 0.2 | 7.2 ± 2.1 | 0.8 |
| Physcion (8) | 0.4 ± 0.0 | 0.3 ± 0.0 | 0.9 | — | — | — |
| Rhein-8-*O*-β-D-glucoside (9) | 0.3 ± 0.0 | 0.3 ± 0.1 | 1.1 | — | — | — |
| Methylisorhein (10) | 1.4 ± 0.1 | 34.0 ± 3.7 | 24.0 | 4.0 ± 1.4 | 10.6 ± 2.3 | 2.6 |
| Aloe-emodin (11) | 37.1 ± 3.6 | 32.4 ± 2.2 | 0.9 | — | — | — |

Antipyrine, high permeability reference compound; atenolol, low permeability reference compound; rhodamine 123, P-gp probe substrate; sulfasalazine, MRP2 probe substrate; estrone-3-sulfate, BCRP probe substrate. Inhibitors, the mixture of verapamil (P-gp inhibitor), MK571 (MRP2 inhibitor), and novobiocin (BCRP inhibitor). An efflux ratio (EfR) greater than 3 was considered as a positive result, suggesting that the investigational compound was an *in vitro* substrate of efflux transporter(s).

In terms of the other anthraquinone constituents with a compound dose of > 0.3 μmol/day, emodin-8-*O*-glucoside (**2**) and physcion (**8**) were found to have poor membrane permeability, which limited their passive intestinal absorption. Although chrysophanol (**1**), chrysophanol-8-*O*-glucoside (**4**), emodin (**5**), physcion-8-*O*-glucoside (**6**), chrysophanol-1-*O*-glucoside (**7**), and aloe-emodin (**11**) showed high or intermediate permeability, **1**, **5**, **6**, and **11** exhibited poor water solubility, which could limit their passive intestinal absorption (Table S5). Additionally, their pre-absorption deglycosylation by human intestinal LPH transforming **4** and **7** into unabsorbable **1** and the efflux action of intestinal apical transporters on them restricted their passive intestinal penetration (Table 4). None of these compounds was a substrate of ASBT or TAUT (Table 3).

### 3.4. Inhibitory activities, against SARS-CoV-2 3CL^pro^, of circulating Dahuang compounds and other circulating compounds of LianhuaQingwen

Rhein (**3**) and methylisorhein (**10**) inhibited 3CL^pro^ of SARS-CoV-2 with IC_50_ values of 4.6 and 37.2 μmol/L, respectively (Fig. 5). However, despite being tested at 100 μmol/L, other Dahuang compounds, such as 4-*O*-methylgallic acid (**M42_M2_**), gallic acid (**42**), and aloe-emodin (**11**), showed less than 50% inhibition of the viral protease. Furthermore, following our prior and ongoing pharmacokinetic investigations on LianhuaQingwen, we singled out several LianhuaQingwen compounds originating from the capsule’s other components and subjected them to SARS-CoV-2 3CL^pro^ inhibition tests. These compounds, unchanged or metabolized, were glycyrrhetic acid, 24-hydroxyglycyrrhetic acid, ephedrine, pseudoephedrine, hydroxytyrosol-3-*O*-sulfate, hydroxytyrosol, phillygenin, phillygenin-*O*-sulfate, pinoresinol, and menthol-1-*O*-glucuronide and showed less than 50% inhibition of the viral protease, despite being tested at 100 μmol/L.

**Fig. 5.**
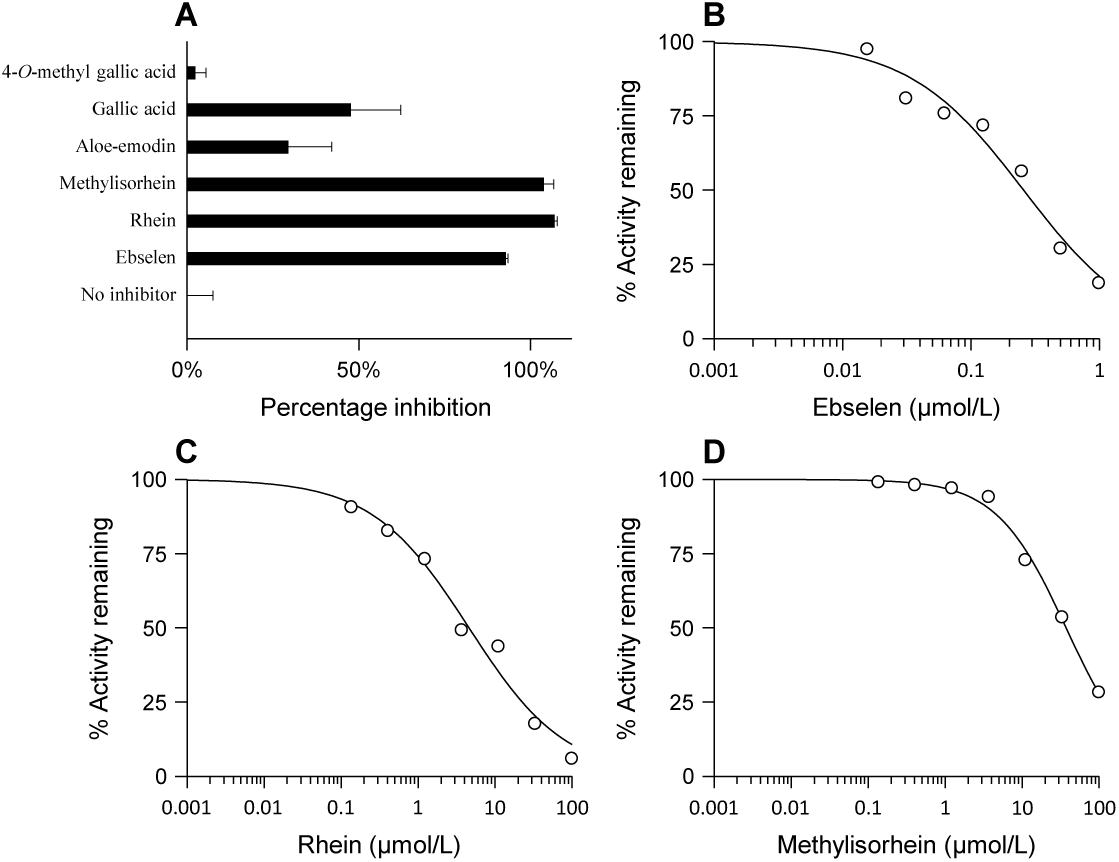
Inhibitory activities against of Dahuang compounds against SARS-CoV-2 3CL^pro^. (A) Percentage inhibition of 3CL^pro^ by circulating Dahuang compounds at 100 μmol/L. (B) IC50 of positive drug ebselen against 3CL^pro^. (C and D) IC50 of rhein (**3**) and methylisorhein (**10**) against 3CL^pro^, respectively.

### 3.5. Expression and cellular localization of Taut and Asbt in rat intestine and lung

The specificity of the anti-TAUT and anti-ASBT antibodies was assessed using Western Blot analysis. HEK-293 cells expressing human TAUT, human ASBT, rat Taut, or rat Asbt with a Flag-tag were used for the experiment. The Flag-tag antibody successfully detected all four plasmids, confirming their expression in the cells. The anti-ASBT antibody recognized both human ASBT and rat Asbt, showing a band at 40 kDa. Similarly, the anti-TAUT antibody detected a band at 65 kDa for both human TAUT and rat Taut (Fig. 6A). These findings validate the reliability of the anti-ASBT and anti-TAUT antibodies in detecting human ASBT/Asbt and TAUT/Taut proteins, respectively. Furthermore, the protein expression levels of Asbt and Taut were examined in rat intestine, liver, and lung tissues using Western Blot analysis with anti-ASBT and anti-TAUT antibodies. Both proteins were detected in the bronchus, lung, liver, and intestine tissues of rats (Fig. 6B). Rat Asbt showed high expression in the bronchus, while Taut exhibited high expression in the liver. Additionally, immunohistochemical analysis was performed on rat lung and intestine tissues. In the bronchial epithelia, rat Asbt was primarily localized on the basolateral membrane, whereas rat Taut was extensively found in the alveolar epithelia and basolateral membrane of bronchial epithelia. Both Asbt and Taut were found in the apical membrane of epithelia in the intestinal villi (Fig. 6C).

**Fig. 6.**
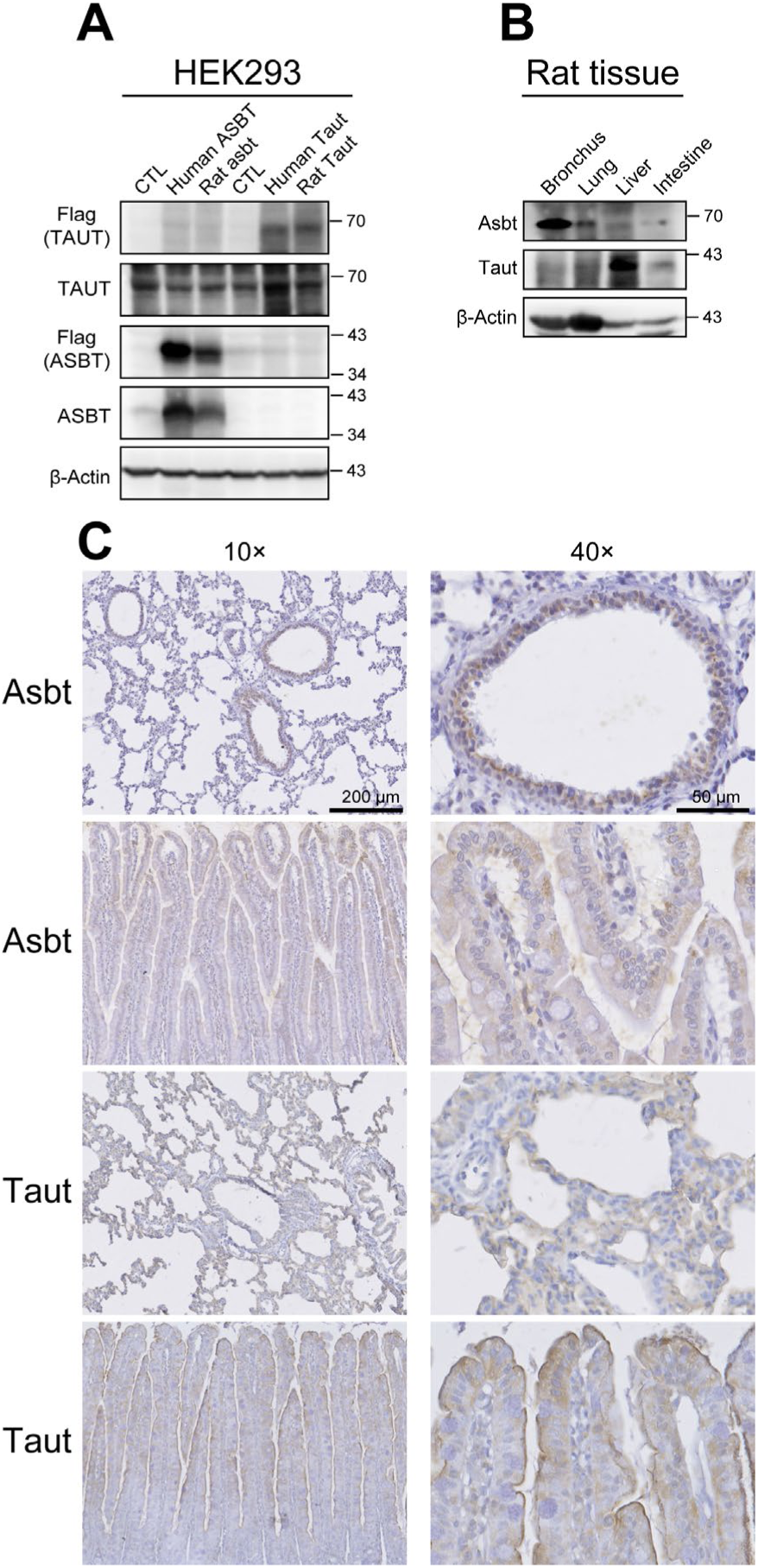
Expression and cellular localization of Taut and Asbt proteins in the rat intestine and lung. (A) Western blot analysis of human ASBT, rat Asbt, human TAUT, or rat Taut proteins in the transfected HEK-293 cells. Representative autoradiographs of the Western blots demonstrating 40-kDa human ASBT and rat Asbt protein bands in the homogenates of human ASBT- and rat Asbt-transfected HEK293 cells, respectively. Similarly, 65-kDa human TAUT and rat Taut proteins are observed in the homogenates of human TAUT- and rat Taut-transfected HEK293 cells, respectively. (B) Western blot analysis of Asbt and Taut proteins in rat lung, liver, and intestine tissues. In the homogenates of these tissues, 40-kDa Asbt protein bands and 65-kDa Taut protein bands are detected in rat bronchus, lung, liver, and intestine. (C) Immunohistochemical localization of Asbt and Taut proteins in rat intestine and lung tissues.

## 4. Discussion

For a complex Chinese herbal medicine, only a few key constituents with drug-like properties are bioavailable at loci responsible for the medicine’s therapeutic action, as opposed to all constituents [21]. Multi-compound pharmacokinetic investigation is a useful approach to accurately identify such potential herbal compounds without any omissions, relying on thorough comprehension of the medicine’s chemical composition [37]. Such bioavailability of LianhuaQingwen compounds was investigated to evaluate the medicine’s efficacy and safety in treating acute viral respiratory illnesses with respect to their systemic exposure and targeted reachability within the body [20]. As part of our multi-compound pharmacokinetic researches on LianhuaQingwen, the previous investigation found that the major circulating Gancao compounds, glycyrrhetic acid and 24-hydroxyglycyrrhetic acid (a new Gancao metabolite), could reach and inhibit renal distal tubular 11β-hydroxysteroid dehydrogenase type 2, a target responsible for Gancao-induced pseudoaldosteronism [20]. In the current investigation, a comprehensive analysis conducted and identified a total of 55 Dahuang constituents (≥0.01 μmol/day) in LianhuaQingwen. However, among these constituents, only three exhibited significant systemic exposure in humans after dosing the capsule. These three constituents were the anthraquinones rhein (**3**) and methylisorhein (**10**; a new Dahuang anthraquinone), and the tannin 4-*O*-methylgallic acid (**M42_M2_**). Interestingly, the levels of the remaining anthraquinones and their metabolites, such as chrysophanol (**1**), emodin (**5**), physcion (**8**), and aloe-emodin (**11**), were either undetectable or found in very low quantities in human plasma. This significant variation in systemic exposure to Dahuang anthraquinones could be attributed to two main factors: transporter-mediated intestinal absorption and LPH-mediated pre-absorption metabolism. Dahuang anthraquinones, including **3** and **10**, have poor water solubility, low membrane permeability, and/or intestinal apical efflux. These properties hindered their passive absorption in the intestine, meaning they could not easily cross the intestinal membrane on their own. However, in the case of **3** and **10**, the human intestinal apical uptake transporters ASBT and TAUT, as well as the human intestinal basolateral efflux transporters MRP1/3/4, could mediate their intestinal absorption and resulted in their significant systemic exposure in human after dosing the capsule. It should be noted that other Dahuang anthraquinones did not exhibit human ASBT- or TAUT-mediated absorption. Additionally, the enzyme LPH present in the human intestinal epithelia played a crucial role in facilitating the transformation of rhein-8-*O*-β-D-glucoside (**9**) into readily absorbable **3**. Interestingly, this enzyme also transformed chrysophanol-8-*O*-β-D-glucoside (**4**) and chrysophanol-1-*O*-β-D-glucoside (**7**) into a non-absorbable **1**, despite their favorable characteristics of being highly water-soluble and easily permeable.

In addition to systemic exposure, targeted reachability of rhein (**3**) and methylisorhein (**10**) was also an important factor for the efficacy of LianhuaQingwen capsule, a medication used in China to treat viral respiratory illnesses, including severe acute respiratory syndrome (SARS), Middle East respiratory syndrome (MERS), and coronavirus disease 2019 (COVID-19). Infections of SARS-CoV, MERS-CoV, and SARS-CoV-2 could cause severe respiratory pathologies and lung injuries by infecting bronchial epithelial cells and pneumocytes in humans [38, 39]. Therefore, it was crucial for the treatment to specifically target the lungs. The 3CL^pro^ enzyme, due to its essential role in viral replication and high degree of conservation across all coronaviruses, as well as its absence in human homologs, has been identified as an attractive target for the development of drug against coronaviruses such as SARS-CoV-2 [38, 40]. Rhein (**3**) and methylisorhein (**10**) demonstrated the ability to inhibit this enzyme, with IC_50_ values of 4.6 and 37.2 μmol/L, respectively (Fig. 5). Like their human counterparts, TAUT and ASBT, mentioned above in the context of intestinal absorption of **3** and **10**, rat Taut and rat Asbt was also found to transport these two compounds (Fig. S2). The mRNA of human TAUT was found to be widely expressed in various human tissues, such as the lungs, kidneys, and liver, due to the recognized role of taurine, an endogenous substrate of TAUT, as an organic osmolyte [41, 42]. In contrast, ASBT mRNA was exclusively detected in the human ileum and kidney [43, 44]. Furthermore, the ASBT protein was found to be predominantly located on the apical membrane of enterocytes in the human terminal ileum [44, 45]. The intestinal TAUT and ASBT was considered a favorite target for developing “drug inhibitor” to regulate the levels of bile acids, cholesterol, lipids, and glucose, or “drug substrate” to improve its oral bioavailability [46, 47]. Although the precise localization of human TAUT and human ASBT proteins in lung epithelia remains unclear, their rat orthologues, Asbt and Taut, was observed in alveolar epithelia and the basolateral membrane of rat bronchial epithelia, as well as the apical membrane of epithelia in the intestinal villi in this investigation (Fig. 6). The uptake of circulating **3** and **10** into lung epithelia could be expected to exert their potential antiviral effects in humans. By targeting the lung epithelia and inhibiting the viral protease enzyme in humans, these two compounds had the potential to combat respiratory illnesses caused by coronaviruses such as SARS-CoV-2. Collectively, systemic exposure to and targeted reachability of **3** and **10** could be significant factors in the efficacy of LianhuaQingwen capsule. Further research on pulmonary localizations and genetic variations of TAUT and ASBT in humans will provide additional insights into the mechanisms underlying the antiviral effects of **3** and **10**.

It is worth noting that systemic exposure to Dahuang anthraquinones and tannins may vary considerably between rats and humans after dosing LianhuaQingwen. While rhein (**3**) and methylisorhein (**10**) were the primary anthraquinones found in both rat and human plasma, rats exhibited additional circulating anthraquinones and their metabolites, including rhein-*O*-glucuronide (**M3_G1_**), that were either undetected or present in limited quantities in human plasma. In humans, the principal circulating compound for tannins was 4-*O*-methylgallic acid (**M42_M2_**), while both **M42_M2_** and gallic acid (**42**) were the major circulating compounds in rats. Glucuronidation was the main metabolic pathway of anthraquinones, including **3**, in rats after dosing a Dahuang extract, while methylation was the main metabolic pathway of gallic acid [48, 49]. Rhein was reported the only compound that humans were significantly exposed to after orally dosing a Dahuang extract [50]. To date, the pharmacokinetic mechanisms governing its significant systemic exposure, including its glucuronidation, remain unclear in humans. Our investigation revealed that, apart from rhein being absorbed in the intestines through human ASBT/TAUT-mediated intestinal uptake from the intestinal lumen and human MRP1/3/4-mediated intestinal efflux into the blood, poor metabolism of **3** in humans was found to be another factor contributing to its significant systemic exposure. This was evident from our observation that there was no significant difference in the systemic exposure to total **3** (including both free and conjugated forms) and free **3** in humans after oral administration of LianhuaQingwen (Fig. S3). This finding was further supported by the observed poor *in vitro* human hepatic glucuronidation, sulfation, and oxidation of **3** (Fig. S1). Methylisorhein (**10**) exhibited a similar situation in humans. The intestinal absorption of **3** and **10** in rats was also mediated by rat orthologues of human ASBT and human TAUT. However, the molecular mechanisms by which other Dahuang anthraquinones and their metabolites were significantly exposed in rats remain unclear. Due to practical and ethical concerns associated with human experimentation, the use of animal models, specifically rats and mice, has become essential in antiviral research. These models are crucial for evaluating the safety and efficacy of antiviral drugs, in addition to conducting in vitro studies on their antiviral activities. Safety concerns arised due to the presence of the quinone moiety in the structure of these anthraquinones. Studies have focused on the potential hepatotoxicity of Dahuang, specifically investigating its anthraquinones and tannins[6]. However, these investigations have primarily relied on preclinical models, especially rat models [6, 51]. Even when assessing the toxicity of specific compounds like **3**, the doses needed to induce toxicity were considerably higher than the typical doses used in clinical settings [51]. Therefore, such interspecies differences in systemic exposure to Dahuang anthraquinones should be taken into account when evaluating the safety and efficacy of the Dahuang-containing herbal medicine in preclinical studies. Further research is needed to fully understand the implications of the interspecies differences associated with molecular mechanisms, in order to accurately assess its potential benefits and risks in humans.

In the field of herbal medicine, anthraquinones are found in a diverse range of plant species, particularly in the families *Polygonaceae* (such as the Chinese medicinal herb Dahuang), *Fabaceae* (Juemingzi and Fanxieye), and *Rubiaceae* (Qiecao). Anthraquinones can be present in these herbs both in their free form as aglycones (like rhein) and bound to sugar residues (like rhein-8-*O*-β-D-glucoside). The quantity of free and glycosylated anthraquinones in herbal medicines can be affected by factors like the specific origin, variety, and processing methods of the component herb. These variations in the active constituents of the herb can affect quality and efficacy of the herbal medicines. For example, in the case of Dahuang, which includes the rhizomes and roots of *R. palmatum* L., *R. tanguticum* Maxim. ex Balf., and *R. officinale* Baill. from the *Polygonaceae* family, the ratio of free and glycosylated rhein content can vary greatly, ranging from 0.001 to 65 [52]. In herbal medicines rich in β-glycosylated anthraquinones, lactase or lactase-phlorizin hydrolase (LPH, EC 3.2.1.23/62) plays a significant role in the hydrolysis of β-glycosylated anthraquinones to free anthraquinones (Fig. 4). This was demonstrated by the significant systemic exposure of free rhein (rather than rhein-8-*O*-β-D-glucoside), with a short *T*_max_ value of 0.5 h, in rats after oral administration of rhein-8-*O*-β-D-glucoside (Fig. S4). LPH is a primary β-glycosidase located in the intestinal microvillus membrane and is responsible for the extracellular hydrolysis of most β-glycosylated xenobiotics, such as glycosylated flavonoids [53]. This hydrolase is predominantly expressed in the jejunum, with lower levels of expression in the proximal and distal ends of the intestines [54]. When the activity of intestinal LPH becomes inhibited or decreased, β-glycosylated anthraquinones reach the colon. It is presumed that the colonic concentration of free anthraquinones increased, resulting in a higher likelihood of inducing local drug effects. This can be attributed to the presence of colonic microbiota and the absence of intestinal uptake transporters, such as ABST [44, 45]. Therefore, by understanding the content and extraction of free and glycosylated anthraquinones in herbal medicines, as well as the role of LPH in their hydrolysis, researchers can better comprehend the potential therapeutic effects and mechanisms of action of these anthraquinone-containing medicinal plants.

## 5. Conclusion

In this investigation, we examined systemic exposure and targeted reachability of Dahuang compounds in humans through a multi-compound pharmacokinetic investigation of a Dahuang-containing medicine LianhuaQingwen. During the investigation, 55 constituents (≥0.01 μmol/day), originating from Dahuang (*R. palmatum* rhizomes and roots), were identified and characterized in LianhuaQingwen. However, only three Dahuang compounds - rhein (**3**), methylisorhein (**10**), and 4-*O*-methylgallic acid (**M42_M2_**) - exhibited significant systemic exposure in humans after dosing the capsule. Two absorption mechanisms for **3** and **10** across the small intestine have been proposed: active intestinal uptake of **3** and **10** by human ASBT/TAUT and human MRP1/3/4, and gut-luminal hydrolysis of rhein-8-*O*-β-D-glucoside (**9**) by LPH, followed by the absorption of released **3** by the intestinal transporters. Another contributing factor to their significant systemic exposure was the poor metabolism of **3** and **10** in humans. In contrast, the other anthraquinones and their metabolites were either undetectable or present in low concentrations in human plasma. This can be attributed to their poor water solubility, low membrane permeability, and/or intestinal apical efflux, which hindered their absorption in the intestines. Additionally, targeted reachability of circulating **3** and **10** could be achieved as rat orthologues of human ASBT/TAUT was observed in alveolar epithelia and basolateral membrane of bronchial epithelia, where coronaviruses could infect and replicate. These two compounds showed potential ability to inhibit the 3CL^pro^ enzyme responsible for viral replication in coronaviruses. This suggested that the compounds had potential use in treating respiratory illnesses caused by coronaviruses. It is important to note that the circulating Dahuang anthraquinones and tannins differed significantly between humans and rats after dosing LianhuaQingwen. This highlights the significance of considering interspecies differences when evaluating the safety and efficacy of the Dahuang-containing medicines. Overall, this investigation, along with similar studies on other components, contributes to defining the therapeutic benefits of Dahuang-containing medicines. It also underscores the need to establish conditions for the safe use of these medicines by considering interspecies differences.

## Data Availability

All data produced in the present study are available upon reasonable request to the authors
All data produced in the present work are contained in the manuscript

## CRediT author statement

**Nan-Nan Tian**: Investigation, Validation, Formal analysis, Visualization; **Ling-Ling Ren**: Investigation, Validation, Formal analysis, Visualization; **Ya-Xuan Zhu**: Investigation, Validation, Formal analysis; **Jing-Ya Sun**: Investigation, Formal analysis, Visualization; **Jun-Lan Lu**: Investigation, Validation; **Jia-Kai Zeng**: Investigation, Validation; **Feng-Qing Wang**: Investigation; **Fei-Fei Du**: Investigation; **Xi-He Yang**: Investigation; **Shu-Ning Ge**: Investigation; **Rui-Min Huang**: Conceptualization, Methodology, Formal analysis; **Wei-Wei Jia**: Investigation, Validation, Formal analysis, Visualization, Conceptualization, Methodology, Writing - Original draft preparation, Reviewing and Editing, Project administration, Funding acquisition; **Chuan Li**: Conceptualization, Methodology, Formal analysis, Writing - Original draft preparation, Reviewing and Editing, Supervision, Funding acquisition.

## Acknowledgements

This work was funded by grants from the National Natural Science Foundation of China (Grant Nos.: 82074176 and 81503345), Innovation Team and Talents Cultivation Program of National Administration of Traditional Chinese Medicine (Grant No.: ZYYCXTD-C-202009), and the National Key R&D Program of China (Grant No.: 2018YFC1704500).

## Appendix A. Supplementary data

## Abbreviations

3CL^pro^: 3-chymotrypsin-like protease
ASBT/Asbt: apical sodium-dependent bile acid transporter
TAUT/Taut: taurine transporter

