## Supplementary data for "Pharmacokinetics-based identification of antiviral compounds of *Rheum palmatum* rhizomes and roots (Dahuang)"

### Original article

### CONTENTS

**Supplementary Materials and methods** 2

**Table S1** Commercially available reference compounds. 8

**Table S2** NMR data of methylisorhein. 9

**Table S3** LianhuaQingwen constituents originating from Dahuang. 10

**Table S4** Unbound fractions of Dahuang compounds in human plasma ($f_u$). 13

**Table S5** Estimation of maximum absorbable dose (MAD) of Dahuang anthraquinones. 14

**Table S6** Net transport ratios for *in vitro* transport of rhein (3) and methylisorhein (10) mediated by various human efflux transporters. 15

**Fig. S1.** *In vitro* metabolism of rhein (**3**) and methylisorhein (**10**) mediated by human enzymes. Glucuronidation of rhein (A) and methylisorhein (B); sulfation of rhein (C) and methylisorhein (D); oxidation of rhein (E) and methylisorhein (F). Solid dots, rhein or methylisorhein; open dots, the respective metabolites. 16

**Fig. S2.** *In vitro* transport of rhein (**3**) and methylisorhein (**10**) mediated by rat transporters. (A) Transport of rhein by rat Taut- and Asbt-transfected HEK-293 cells. (B) Transport of methylisorhein by rat Taut- and Asbt-transfected HEK-293 cells. MC, mock HEK-293 cells.

**Fig. S3.** Comparison of area under plasma concentration-time curve (AUC) of free and total rhein (**3**, A) or methylisorhein (**10**, B) in humans after oral administration of LianhuaQingwen at 12 capsules/person. The plasma samples underwent pre- and post-hydrolysis treatments with hydrochloric acid (4 M) at 80°C for 30 min to release free anthraquinone from its various forms, including glycosylated, glucuronidated, and sulfated forms.

**Fig. S4.** Plasma concentration-time profiles of rhein (**3**) and rhein-8-*O*-β-D-glucoside (**9**) in rats after oral administration of rhein-8-*O*-β-D-glucoside at 1.8 mg/kg. Solid dots, rhein; Open dots, rhein-8-*O*-β-D-glucoside.

### Supplementary Materials and methods

#### *2.1. Study materials*

LianhuaQingwen capsules are a type of hard gelatin capsule produced by Shijiazhuang Yiling Pharmaceutical Co., Ltd (Yiling Pharmaceutical; Shijiazhuang, Hebei Province, China) and have a Chinese NMPA drug ratification number of GuoYaoZhunZi-Z20040063. Each capsule contained 0.35 g herbal extract, which was prepared from a combination of 0.255 g of Lianqiao (*F. suspensa* fruits), 0.255 g of Jinyinhua (*L. japonica* flower buds), 0.085 g of stir-fried Mahuang with honey (processed *E. sinica* stems), 0.085 g of stir-fired Kuxinren (processed *P. sibirica* seeds), 0.255 g of Shigao (*Gypsum Fibrosum*), 0.255 g of Banlangen (*I. indigotica* roots), 0.255 g of Mianmaguanzhong (*D. crassirhizoma* rhizome), 0.255 g of Yuxingcao (*H. cordata* whole plants), 0.085 g of Guanghuoxiang (*P. cablin* overground portion), 0.051 g of Dahuang (*R. palmatum* rhizomes and roots), 0.085 g of Hongjingtian (*R. crenulata* rhizomes and roots), 0.0075 g of Bohenao (menthol), and 0.085 g of Gancao (*G. uralensis* roots). The herb-to-extract ratio is 5.8:1, and the capsule is standardized to contain at least 0.17 mg forsythin. To analyze the capsule's chemical composition and lot-to-lot variability, samples of 14 lots of LianhuaQingwen were obtained from Yiling Pharmaceutical along with samples of individual herbal components, such as Dahuang (*R. palmatum* rhizomes and roots). All these samples were stored at −20°C until analysis.

Purified compounds found in *Rheum* species were obtained from Chroma-Biotechnology (Chengdu, Sichuan Province, China) and Yuanye Bio-Technology (Shanghai, China) (Table S1). One such compound, 3,8-dihydroxy-1-methylanthraquinone-2-carboxylic acid (methylisorhein), was extracted from Dahuang (*R. palmatum* rhizomes and roots) with 75% ethanol. The extract was then fractioned using macroporous resins D101 column-based chromatography and reversed phase Agilent Zorbax SB-C18 5-μm column-based chromatography. The resulting fraction containing methylisorhein was concentrated under reduced pressure and recrystallized to obtain the pure compound. To elucidate the chemical structure of the isolated compound, various nuclear magnetic resonance (NMR) techniques were employed and $^{1}$H NMR, $^{13}$C NMR, DEPT135, $^{1}$H-$^{1}$H COSY, $^{1}$H-$^{13}$C HSQC, and $^{1}$H-$^{13}$C HMBC NMR data were obtained on both Bruker AVANCE III 500 and Bruker AVANCE III HD 600 MHz NMR spectrometers (Table S2).

Caco-2 cells and HEK-293 cells were obtained from American Type Culture Collection (Manassas, VA, USA). Various transporter cDNAs, including human apical sodium-dependent bile acid transporter (ASBT; GeneBank accession number, NM_000452), taurine transporter (TAUT; NM_003043), organic anion-transporting polypeptide 2B1 (OATP2B1; NM_007256), peptide transporter 1 (PEPT1) (NM_005073), PEPT2 (NM_021082), organic anion transporter 1 (OAT1) (NM_004790), OAT2 (NM_006672), OAT3 (NM_004254), OAT4 (NM_018484), and organic cation transporter 2 (OCT2; NM_003058), along with enzyme cDNA lactase-phlorizin hydrolase (LPH; NM_002299), were synthesized and subcloned into pcDNA 3.1(+) expression vectors by Invitrogen Life Technologies (Shanghai, China). In addition, inside-out membrane vesicle suspensions expressing human multidrug resistance protein 1 (MDR1), breast cancer resistance protein (BCRP), multidrug resistance-associated protein 1 (MRP1), MRP2, MRP3, and MRP4 were obtained from Genomembrane (Kanazawa, Japan). Pooled human liver microsomes (HLM) and pooled human liver cytosol (HLC) were obtained from Corning Gentest (Woburn, MA, USA).

Taurocholic acid and [$D_4$]-taurine were obtained from Shanghai Zzbio (Shanghai, China), while other chemicals, including estrone-3-sulfate, glycylsarcosine, *para*-aminohippuric acid, prostaglandin $F_{2a}$, tetraethylammonium, atenolol, antipyrine, rhodamine123, sulfasalazine, glycyrrhizin, estradiol-17β-D-glucuronide, verapamil, MK-571, novobiocin, phlorizin, phloretin, chrysin, 7-hydroxyflavone, and midazolam, were obtained from J&K Scientific (Beijing, China). HPLC-grade methanol, formic acid, ethyl acetate, dimethyl sulfoxide, and deuterated dimethyl sulfoxide were obtained from Sigma-Aldrich (St. Louis, MO, USA).

#### *2.2. Human pharmacokinetic study*

A single-center, open-label human pharmacokinetic study was conducted at Hebei Yiling Hospital (Shijiazhuang, China) to investigate the pharmacokinetics of LianhuaQingwen capsule. The study, which was approved by the hospital's ethics committee and registered with the Chinese Clinical Trials Registry (ChiCTR1900021460), adhered to the Declaration of Helsinki. A total of 14 healthy volunteers (eight men and six women, aged between 19 and 42 years with BMIs ranging between 20.7

and 25.1 kg/m$^2$) provided written informed consent and participated in the study. The details of the human study are described previously [1]. In brief, the participants orally received a single dose of 12 capsules (label daily dose) of LianhuaQingwen (A1802055) on day 1 and days 3–8. On day 1, serial blood samples were collected before and 10 and 30 min and 1, 3, 6, 9, 12, 16, 24, 28, 34, and 48 h after dosing, and serial urine samples were also collected within 0–4, 4–10, 10–24, 24–32, and 32–48 h periods after dosing. On days 3–7, blood samplings were performed before and 12 h after dosing. On day 8, serial blood and urine samples were collected according to the respective time schedules on day 1. The blood samples were heparinized and centrifuged to yield the respective plasma samples. All the human samples were stored at −70°C pending analysis by liquid chromatography/mass spectrometry.

#### *2.3. Supportive in vitro transport and metabolism studies*

##### *2.3.1. Transport study using Caco-2 cell monolayers*

Eleven test Dahuang compounds, including chrysophanol (**1**; 5 μmol/L), emodin-8-*O*-β-D-glucoside (**2**; 50 μmol/L), rhein (**3**; 5 μmol/L), chrysophanol-8-*O*-β-D-glucoside (**4**; 50 μmol/L), emodin (**5**; 5 μmol/L), physcion-8-*O*-β-D-glucoside (**6**; 50 μmol/L), chrysophanol-1-*O*-β-D-glucoside (**7**; 50 μmol/L), physcion (**8**; 5 μmol/L), rhein-8-*O*-β-D-glucoside (**9**; 50 μmol/L), methylisorhein (**10**; 5 μmol/L), and aloe-emodin (**11**; 5 μmol/L), were investigated for their membrane permeability in Caco-2 cell monolayers. The Caco-2 cells (passages 33 and 34) were cultured at a density of approximately 30,000 cells/cm² in Dulbecco's modified Eagle's medium containing 10% fetal bovine serum. The transepithelial electrical resistance of the Caco-2 cell monolayers was measured before each experiment to ensure its integrity, which should be ≥ 400 Ω·cm². The apparent permeability coefficient ($P_{app}$) of test compound is estimated using equation 1:

$$P_{app} = (\Delta Q/\Delta t)/(A \times C_0) \quad (1)$$

where $\Delta Q/\Delta t$ is the linear appearance rate of the test compound on the receiver side in μmol/s, $A$ is the surface area of the cell monolayer (i.e., 0.6 cm$^2$), and $C_0$ is the initial concentration of the test compound on the donor side in μmol/L. Test compounds were classified into the categories of low, intermediate, or high membrane permeability based on their $P_{app}$ values: < 1 × 10$^{-6}$, 1–10 × 10$^{-6}$, or > 10 × 10$^{-6}$ cm/s, respectively [2]. The efflux ratio (EfR) was calculated as the ratio of $P_{app(B\text{-}A)}$ to $P_{app(A\text{-}B)}$ and used to implicate the possible involvement of a transporter-mediated efflux mechanism and a positive outcome was determined with an EfR value > 3 and a statistically significant difference between $P_{app(B\text{-}A)}$ and $P_{app(A\text{-}B)}$. Verapamil, MK571, and novobiocin were added at 100 μmol/L each to inhibit the ABC transporters MDR1, MRP2, and BCRP, respectively, to prevent potential influence of active efflux transport on $P_{app(A\text{-}B)}$. Atenolol and antipyrine were used as low and high permeability reference compounds, respectively, while rhodamine 123, sulfasalazine, and estrone-3-sulfate were used as positive substrates of MDR1, MRP2, and BCRP, respectively. The cell culture and bidirectional transport experimental details are described previously [3].

##### *2.3.2. Transport studies using cells transfected with SLC transporters or membrane vesicles expressing ABC transporters*

Methods for culturing cells and transfecting HEK-293 cells with various human SLC transporters are described previously [4, 5]. To ensure functionality, transfected cells (TCs) and mock cells (MCs) were characterized by measuring cellular uptake of different positive compounds, including: taurocholic acid (for human ASBT), [D$_4$]-taurine (human TAUT), estrone-3-sulfate (human OATP2B1, OAT3, and OAT4), tetraethylammonium (human OCT2), glycylsarcosine (human PEPT1 and PEPT2), *para*-aminohippuric acid (human OAT1), and prostaglandin F2a (human OAT2). The test compounds used for the uptake measurements were: chrysophanol (**1**; 5 μmol/L), emodin-8-*O*-β-D-glucoside (**2**; 50 μmol/L), rhein (**3**; 5 μmol/L), chrysophanol-8-*O*-β-D-glucoside (**4**; 50 μmol/L), emodin (**5**; 5 μmol/L), physcion-8-*O*-β-D-glucoside (**6**; 50 μmol/L), chrysophanol-1-*O*-β-D-glucoside (**7**; 50 μmol/L), physcion (**8**; 5 μmol/L), rhein-8-*O*-β-D-glucoside (**9**; 50 μmol/L), methylisorhein (**10**; 5 μmol/L), aloe-emodin (**11**; 5 μmol/L), gallic acid (**42**; 5 μmol/L), and 4-*O*-methylgallic acid (**M42**$_{\mathbf{M2}}$; 5 μmol/L). The transport rate of the test compounds was estimated using equation 2:

$$\text{Transport} = (C_L \times V_L)/T/W_L \quad (2)$$

where $C_L$ is the concentration of the test compound in the cellular lysate in μmol/L, $V_L$ is the volume of the lysate in μL, $T$ is the incubation time (i.e., 10 min), and $W_L$ is the protein amounts measured in the lysate in mg. Differential uptake between the TC and MC was defined as a net transport ratio (Transport$_{TC}$/Transport$_{MC}$ ratio), where Transport$_{TC}$ and Transport$_{MC}$ are the transport rates of the test compound in the TC and MC, respectively. A net transport ratio >3, with a statistically significant difference between Transport$_{TC}$ and Transport$_{MC}$, indicated a positive result in this laboratory based on previous validation on positive and negative control compounds.

Inside-out membrane vesicles expressing one of the human ABC transporters MDR1, BCRP, MRP1, MRP2, MRP3, and MRP4 were used to assess the transport of rhein (**3**; 5 μmol/L) and methylisorhein (**10**; 5 μmol/L) using a rapid filtration method described previously [4, 5]. Before use, the membrane vesicles were functionally characterized by measuring vesicular transport of glycyrrhizin (a positive substrate of MDR1 and BCRP) and estradiol-17β-D-glucuronide (MRP1, MRP2, MRP3, and MRP4). The transport rate of test compound in pmol/min/mg protein was calculated using the equation 3:

$$\text{Transport} = (C_V \times V_V)/T/W_V \qquad (3)$$

where $C_V$ is the concentration of the compound in the vesicular lysate supernatant in μmol/L, $V_V$ is the volume of the lysate in μL, $T$ is the incubation time (i.e., 10 min) and $W_V$ is the amount of vesicle protein amount per well (i.e., 0.05 mg). Positive results for ATP-dependent transport were defined as a net transport ratio ($\text{Transport}_{ATP}/\text{Transport}_{AMP}$ ratio) >2, with a statistically significant difference between $\text{Transport}_{ATP}$ and $\text{Transport}_{AMP}$.

*2.3.3. Metabolism studies*

LPH plasmids were introduced into the HEK-293 cells with Lipofectamine 2000 transfection reagent (Invitrogen; Carlsbad, CA, USA) and the transiently transfected cells were lysed by freeze-thawing. Before use, the cell lysate was functionally characterized for expressing of human LPH by measuring deglycosylation of phlorizin (a positive substrate of LPH). The test compounds emodin-8-*O*-β-D-glucoside (**2**; 1 μmol/L), chrysophanol-8-*O*-β-D-glucoside (**4**; 1 μmol/L), physcion-8-*O*-β-D-glucoside (**6**; 1 μmol/L), chrysophanol-1-*O*-β-D-glucoside (**7**; 1 μmol/L), and rhein-8-*O*-β-D-glucoside (**9**; 1 μmol/L) were assessed for deglycosylation using the cell lysate. Incubation was maintained at 37 °C for 60 min and samples were taken at 15, 30, and 60 min for evaluation.

Before use, HLM were functionally characterized for expressing of UGT enzymes by measuring glucuronidation of chrysin (a positive substrate of UGTs) and for expressing of P450 enzymes by measuring oxidation of midazolam (a positive substrate of P450s), while HLC was characterized for expressing of SULT enzymes by measuring sulfation of 7-hydroxyflavone (a positive substrate of SULTs). The test compounds rhein (**3**; 1 μmol/L) and methylisorhein (**10**; 1 μmol/L) were assessed for glucuronidation using UDPGA-fortified HLM, sulfation using PAPS-fortified HLC, and oxidation of NADPH-fortified HLM. The detailed incubation conditions were described previously [6] and sampling were performed 60 and 120 min after initiating incubation.

*2.4. Supportive rat pharmacokinetic studies*

Rat studies were conducted at Laboratory Animal Center of Shanghai Institute of Materia Medica (Shanghai, China). The Institutional Animal Care and Use Committee approved the study protocol, which was conducted in adherence with the Guidelines for Ethical Treatment of Laboratory Animals by the Ministry of Science and Technology of China in 2006. Male Sprague-Dawley rats (0.23–0.28 kg) were obtained from Sino-British SIPPR/BK Laboratory Animal (Shanghai, China), housed under specific-pathogen-free conditions at 20–24 °C and relative humidity of 30%–70% with a 12-h light/dark cycle. The rats were given ad libitum access to commercial rat chow and filtered tap water and acclimated to the facilities and environment for three days before use. Femoral artery cannulation or bile duct cannulation was performed on the rats to collect blood or bile samples, respectively, as previously described [7]. Afterward, the rats were housed singly and allowed to recover their preoperative body weights. $CO_2$ was used to euthanize all rats. In total, 18 rats were used in the following three rat studies.

In the first rat study, six rats were given an oral dose of LianhuaQingwen at 3.78 g/kg (nine times the dose translated from the human label daily dose of the capsule 4.2 g/day, using a body surface area normalization method [8]). Serial blood samples (150 μL each) were collected before and 5, 15, and 30 min and 1, 2, 4, 6, 8, 11, 15, 20, and 24 h after dosing, treated with heparin, and centrifuged to yield plasma samples. In the second rat study, six rats, housed singly in rat metabolic cages (with urine collection tubes frozen at −15 °C), were given an oral dose of LianhuaQingwen at 3.78 g/kg via gavage. Serial urine samples were collected within 0–8, 8–24, 24–32, and 32–48 h periods after dosing. In the third rat study, six rats were given an oral dose of LianhuaQingwen at 3.78 g/kg via gavage. Serial bile samples were collected within 0–1, 1–2, 2–4, 4–6, 6–8, 8–10, 10–24, 24–34, and 34–48 h periods after dosing. All the rat samples were stored at −70°C and were subsequently analyzed by liquid chromatography/mass spectrometry.

*2.5. Assay for composition analysis of Dahuang constituents in LianhuaQingwen*

To analyze the composition of LianhuaQingwen, both samples of its component herb Dahuang and itself were extracted with 70% methanol and centrifuged. The supernatants underwent analysis on a Waters Synapt G2 high definition time-of-flight mass spectrometer (Manchester, UK) interfaced via a LockSpray source with a Waters Acquity ultra performance liquid chromatographic separation module (Milford, MA, USA). Chromatographic separation was achieved on a Waters Acquity UPLC BEH 1.7-μm C18 column (100 × 2.1 mm i.d.; Dublin, Ireland; at 45°C) with a mobile phase consisting of solvent A (water/methanol, 99:1, v/v; containing 25 mmol/L formic acid) and solvent B (water/methanol, 1:99, v/v; containing 25 mmol/L formic acid) at a flow rate of 0.3 mL/min using a 42-min gradient (0–2 min at 2% solvent B, 2–32 min from 2% to 98% solvent B, 32–37 min at 98% solvent B, and 37–42 min at 2% solvent B). The mass spectrometer was operated in the sensitive mode at a resolving power of approximately 10,000. The LockSpray source worked in the negative ion mode, the mass spectrometer was externally calibrated over a range of *m/z* 50–1500 using a 5 mmol/L sodium formate solution at 10 μL/min, and mass shifts during acquisition were corrected using leucine enkephalin (*m/z* 554.2615) as a lockmass. The acquisition of $MS^E$ data (in centroid mode, m/z 50–1500) was performed using a trap collision energy of 4 V and a trap collision energy ramp of 20–40 V (sampling cone, 25 V) with a scan time of 0.3 second. The $MS^E$ acquisition time was set over a retention time range of 1–42 min.

In the composition analysis of LianhuaQingwen, detection of constituents originating from Dahuang was achieved using the literature-mined information on candidate constituents (summarized in a compound list containing compounds' names, molecular formulas, molecular masses, and structures). Each candidate constituent detected in LianhuaQingwen was checked for its component origin and the analysis of the components was performed under the same conditions for composition analysis of the medicine. The detected candidate constituents were characterized by comparison of them with their respective reference compounds (when available) with respect to chromatographic retention time, accurate molecular mass/ionization pattern, and fragmentation profile. When the reference compounds were not available, the characterization was achieved by comparison with the respective literature-mined data, including chromatographic elution order with one or more characterized coexisting constituents and mass spectrometry data. Quantification of the characterized constituents for their content levels in LianhuaQingwen was achieved by calibration using respective reference compounds (when available) or using respective structurally similar reference compounds.

*2.6. Bioanalytical assays for analyses of Dahuang compounds and other study compounds*

Two types of bioanalysis were conducted to investigate the pharmacokinetics of Dahuang compounds in LianhuaQingwen administration. The first type involved profiling unchanged and metabolized Dahuang compounds in human/rat samples prepared with two volumes of methanol. The second type involved quantifying specific Dahuang compounds in human/rat samples prepared by acidification with hydrochloric acid and extraction with methanol and *in vitro* samples prepared using two volumes of ice-cold methanol.

Profiling of Dahuang compounds in human/rat study samples was performed using the Acquity ultra performance liquid chromatographic separation module/Waters Synapt G2 mass spectrometer. The profiling entailed detection, characterization, and quantification of the compounds. Both chromatographic separation and mass spectrometry for the compound-profiling analysis were achieved under the same conditions as the preceding composition analysis. Detection and characterization of unchanged Dahuang compounds in the biological samples was achieved using the characterized LianhuaQingwen sample as reference standard. Meanwhile, detection of metabolized Dahuang compounds mainly focused on those derived from the major Dahuang constituents of LianhuaQingwen, and a list of candidate metabolites guiding detection was generated using an Accelrysmetabolite database (version 2015.1; San Diego, CA, USA) [11]. Quantification of these Dahuang compounds in biological matrices was achieved by calibration using respective reference compounds (when available) or using a structurally similar reference compound.

Quantification of specific Dahuang compounds in biological matrices was performed using an Applied Biosystems Sciex Triple Quad 5500 mass spectrometer (Toronto, Ontario, Canada) interfaced via a Turbo V ion source with an Agilent 1290 Infinity II LC system (Waldbronn, Germany). Chromatographic separation was achieved on an Agilent Poroshell 120 EC-C18 column (50 × 2.1 mm, 2.7 μm; Chadds Ford, PA, USA) with a mobile phase consisting of solvent A (water/acetonitrile, 99:1, v/v; containing 10 mmol/L ammonium formate) and solvent B (water/acetonitrile, 1:99, v/v; containing

10 mmol/L ammonium formate) at a flow rate of 0.3 mL/min using a 10-min gradient (0–0.5 min at 1% solvent B, 0.5–3.5 min from 1% to 8% solvent B, 3.5–7.5 min from 8% to 60% solvent B, 7.6–9.0 min from 60% to 98% solvent B, 9–9.1 min from 98% to 1% solvent B, and 9.1–10 min at 1% solvent B). Matrix-matched calibration curves were constructed with reference standards, using weighted ($1/X$) linear regression of the peak areas ($Y$) of the analytes against the corresponding nominal analytes' concentrations ($X$; 7.81, 15.6, 31.3, 62.5, 125, 250, 500, and 1000 nmol/L), and the curves showed great linearity ($R \geq 0.99$). Although an internal standard was not used, the implemented assay validation, following the European Medicines Agency Guideline on bioanalytical method validation (2012; www.ema.europa.eu), confirmed that the developed assays were reliable and reproducible for the intended use. The lower limits of quantification of the assays were between 7.81–15.6 nmol/L, and the upper limits of quantification were 1000 nmol/L. The intra-batch accuracy and precision were within 89.4%–106% and 2.91%–12.6%, respectively, while inter-batch values were within 92.7%–107% and 5.46%–14.0%, respectively. The coefficients of variation reflecting matrix effects on assays were 2.76–12.4%. The analyte stability was also evaluated under conditions mimicking the analytical process, including storage at 24°C for 4 h and storage at 8°C for 24 h, with the analytes remaining stable under the test conditions. All the validation results were within the acceptable ranges.

Concentrations of various compounds in *in vitro* study samples were analyzed using liquid chromatography/mass spectrometry. The compounds included taurocholic acid, [$D_4$]-taurine, estrone-3-sulfate, glycylsarcosine, *para*-aminohippuric acid, prostaglandin $F_{2a}$, tetraethylammonium, antenolol, antipyrine, rhodamine 123, sulfasalazine, phlorizin, phloretin, chrysin-7-*O*-glucuronide, flavone-7-*O*-sulfate, midazolam-1'-hydroxylation, as well as Dahuang compounds.

#### *2.7. In vitro assessment of SARS-CoV-2 3CL$^{pro}$ protease inhibition*

The proteolytic activity of the 3-chymotrypsin-like protease (3CL$^{pro}$) of SARS-CoV-2 was monitored with a fluorescence resonance energy transfer (FRET) assay. The activity of recombinant SARS-CoV-2 3CL$^{pro}$ was evaluated in cleaving a synthetic fluorogenic peptide substrate MCA-AVLQSGFR-Lys(Dnp)-Lys-$NH_2$, which was derived from the N-terminal auto-cleavage sequence from the viral protease. The cleaved MCA peptide fluorescence was measured by a SpectraMax M2 microplate reader (Molecular Devices; CA, USA) at an excitation wavelength of 325 nm and an emission wavelength of 393 nm. The assay reaction buffer contained 50 mM Tris-HCl (pH 7.3), 1 mM EDTA, and 20 µM peptide substrate. Enzymatic reactions were initiated with the addition of 100 nM SARS-CoV-2 3CL$^{pro}$ and allowed to proceed at 37 °C for 10 min (under linear cleavage condition). The Michaelis-Menten constant of the peptide substrate for SARS-CoV-2 3CL$^{pro}$ was measured to determine the substrate concentration used in the following inhibition assessment. Ebselen, a positive inhibitor of SARS-CoV-2 3CL$^{pro}$, decreased the fluorescence. The inhibition potencies of the test Dahuang compounds, such as rhein (**3**), methylisorhein (**10**), aloe-emodin (**11**), 4-*O*-methylgallic acid (**M42$_{M2}$**), and gallic acid (**42**), were initially screened at a concentration of 100 µmol/L. The Dahuang compounds that showed ≥ 50% inhibition in the initial screening were further evaluated at multiple concentrations for their half-maximal inhibitory concentrations ($IC_{50}$) for SARS-CoV-2 3CL$^{pro}$. For the $IC_{50}$ assessment, the recombinant protease was incubated with the peptide substrate at 1.4 µmol/L (the $K_m$ for SARS-CoV-2 3CL$^{pro}$) in the presence or absence of the test Dahuang compound.

#### *2.8. Western blot*

HEK-293 cells were transfected with empty vector, human Flag-ASTB, human Flag-TAUT, rat Flag-ASBT, or rat Flag-TAUT plasmids for 48 hours. After that, the cells were harvested and lysed with RIPA lysis buffer (pH 7.5) containing 150 mM NaCl, 1 mM $Na_3VO_4$, 50 mM Tris-HCl, 25 mM NaF, 1% NP-40, 0.5% sodium deoxycholate, 0.1% SDS, and 1% phosphatase inhibitor cocktails. Rat lung, bronchus, liver, and intestine were collected and lysed in the same RIPA lysis buffer using a grinder. Subsequently, 20 µg of the aforementioned proteins were loaded onto SDS-PAGE and transferred to nitrocellulose membranes (Millipore, Billerica, MA, USA). The membranes were then incubated overnight at 4°C with primary antibodies against ASBT (1:1000, 25245-1-AP; Proteintech, Wuhan, Hubei Province, China) or TAUT (1:1000, PA5-98161; Invitrogen, Waltham, MA, USA). For detection, the membranes were treated with secondary antibodies conjugated to horseradish peroxidase (Jackson, West Grove, PA, USA) and the immunoreactive proteins were visualized using the West Pico PLUS Chemiluminescent Substrate (Thermo, Waltham, MA, USA) and imaged using chemiluminescent imaging.

*2.9. Immunohistochemistry*

Rat lung and intestine tissues were fixed in 4% paraformaldehyde and embedded in paraffin. The tissues were then sectioned into 5 μm thick paraffin sections and deparaffinized. Antigen retrieval was performed using Tris-EDTA (pH 9.0). For immunohistochemistry staining, the sections were incubated with primary antibodies against ASBT (1:100, AB203205; Abcam, Cambridge, UK) or TAUT (1:500, PA5-98161; Invitrogen, Waltham, MA, USA), followed by incubation with biotinylated goat anti-rabbit IgG as secondary antibodies. Signals were amplified using a DAB kit (Maixin Bio, Fuzhou, Fujian Province, China) and counterstained with hematoxylin.

*2.10. Data Processing*

After composition analysis, all the detected and characterized Dahuang constituents were ranked in descending order according to their respective daily doses (compound doses) and graded into different levels, i.e., 1–10, 0.1–1, 0.01–0.1, and < 0.01 μmol/day. The daily compound dose was calculated by multiplying the compound level in LianhuaQingwen and the capsule's label daily dose of 4.2 g/day.

Pharmacokinetic parameters of Dahuang compounds were estimated by noncompartmental analysis using Kinetica (version 5.0; Thermo Scientific, Philadelphia, PA, USA). The renal clearance ratio ($R_{rc}$) was calculated using equation 4:

$$R_{rc} = CL_R/(GFR \times f_u) \quad (4)$$

where $CL_R$ is the renal clearance, GFR is the glomerular filtration rate in humans (107 mL/h/kg) [9], and $f_u$ is the unbound fraction in human plasma.

All data are expressed as the mean ± standard deviation. Statistical analysis was performed by two-tailed unpaired Student t-test using SPSS Statistics software (version 19.0; IBM, Chicago, IL, USA). A value of $P < 0.05$ was considered to be the minimum level of statistical significance.

**Table S1** Commercially available reference compounds.

| Compound | Purity | CAS number | Source |
|---|---|---|---|
| *Anthraquinones* | | | |
| Chrysophanol | ≥98% | 481-74-3 | Chroma-Biotechnology |
| Emodin | ≥98% | 518-82-1 | (Chengdu, Sichuan Province, China) |
| Rhein | ≥98% | 478-43-3 | |
| Aloe-emodin | ≥98% | 481-72-1 | |
| Physcion | ≥98% | 521-61-9 | |
| Chrysophanol-1-*O*-β-D-glucoside | ≥97% | 4839-60-5 | |
| Chrysophanol-8-*O*-β-D-glucoside | ≥98% | 13241-28-6 | |
| Emodin-1-*O*-β-D-glucoside | ≥98% | 38840-23-2 | |
| Emodin-8-*O*-β-D-glucoside | ≥98% | 23313-21-5 | |
| Rhein-8-*O*-β-D-glucoside | ≥98% | 113443-70-2 | |
| Aloe emodin-8-*O*-β-D-glucoside | ≥98% | 33037-46-6 | |
| Physcion-8-*O*-β-D-glucoside | ≥98% | 26296-54-8 | |
| *Anthrones* | | | |
| Sennoside A | ≥98% | 81-27-6 | Chroma-Biotechnology |
| *Galloylglucoses* | | | |
| Gallic acid | ≥98% | 149-91-7 | Yuanye Biotechnology |
| 3-*O*-methylgallic acid | ≥98% | 3934-84-7 | (Shanghai, China) |
| 4-*O*-methylgallic acid | ≥95% | 4319-02-2 | |
| *Flavonoids* | | | |
| (+)-Catechin | ≥98% | 154-23-4 | Yuanye Biotechnology |
| (–)-Epigallocatechin-3-*O*-gallate | ≥98% | 989-51-5 | Chroma-Biotechnology |
| Vitexin | ≥98% | 3681-93-4 | |
| Eriodictyol | ≥98% | 552-58-9 | |
| *Naphthalenes* | | | |
| Rheumone A | ≥98% | 1415-73-2 | Chroma-Biotechnology |
| Torachrysone-8-*O*-β-D-glucoside | ≥98% | 64032-49-1 | |
| *Stilbenes* | | | |
| Rresveratrol | ≥98% | 501-36-0 | Chroma-Biotechnology |
| Rhapontigenin | ≥98% | 500-65-2 | |
| *Pyranones* | | | |
| 2,5-Dimethyl-7-hydroxychromone | ≥98% | 38412-47-4 | Chroma-Biotechnology |
| 2-Methyl-5-acetonyl-7-hydroxychromone | ≥95% | 28955-30-8 | |
| *Phenylbutanones* | | | |
| Lindleyin | ≥98% | 59282-56-3 | Chroma-Biotechnology |
| Isolindleyin | ≥98% | 87075-18-1 | |

**Table S2** NMR data of methylisorhein.

| Position | $^1$H NMR | $^{13}$C NMR | $^1$H NMR [multiplicity (J in Hz)] |
|---|---|---|---|
| 1 | | 141.1 | |
| 2 | | 122.3 | |
| 3 | | 159.6 | |
| 4 | 7.57 | 112.3 | s |
| 5 | 7.62 | 118.3 | dd, J=7.5, 1.2 |
| 6 | 7.71 | 136.0 | dd, J=8.3, 7.5 |
| 7 | 7.32 | 124.4 | dd, J=8.3, 1.2 |
| 8 | | 161.4 | |
| 9 | | 189.3 | |
| 10 | | 182.0 | |
| 11 | 2.7 | 19.8 | s |
| 12 | | 168.2 | |
| 4a | | 136.3 | |
| 8a | | 116.8 | |
| 9a | | 130.9 | |
| 10a | | 132.4 | |
| 8-OH | 12.88 | | s |

s, singlet; d, double.

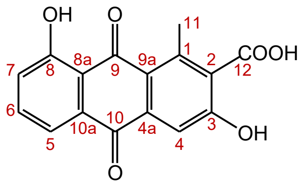

**Table S3** LianhuaQingwen constituents originating from Dahuang.

| | | Liquid chromatography/mass spectrometry data | | | | | Molecular | |
|---|---|---|---|---|---|---|---|---|
| ID | Constituent | $t_R$ (min) | $[M–H]^-$ (*m/z*) | Mass error (ppm) | Fragmentation profile (*m/z*) | Molecular formula | mass (Da) | Dose level (RSD) (μmol/person) |
| *Anthraquinones* | | | | | | | | |
| **1** | Chrysophanol | 26.32 | 253.0511 | 4.0 | 225.0569, 183.0091 | $C_{15}H_{10}O_4$ | 254.0579 | 2.23 ± 0.22 (9.96%) |
| **2** | Emodin-8-*O*-β-D-glucoside | 18.94 | 431.0966 | −2.8 | 269.0443, 225.1192 | $C_{21}H_{20}O_{10}$ | 432.1056 | 1.47 ± 0.14 (9.33%) |
| **3** | Rhein | 22.98 | 283.0247 | 1.4 | 239.0348, 211.0422, 183.0411 | $C_{15}H_8O_6$ | 284.0321 | 1.45 ± 0.20 (13.83%) |
| **4** | Chrysophanol-8-*O*-β-D-glucoside | 18.62 | 415.1031 | 0.5 | 277.0516, 253.0507, 225.0460 | $C_{21}H_{20}O_9$ | 416.1107 | 1.36 ± 0.17 (12.14%) |
| **5** | Emodin | 25.61 | 269.0455 | 1.9 | 241.0667, 225.0515, 197.0307 | $C_{15}H_{10}O_5$ | 270.0528 | 1.13 ± 0.09 (8.31%) |
| **6** | Physcion-8-*O*-β-D-glucoside | 20.53 | 445.1133 | −0.4 | 283.0608, 240.0454 | $C_{22}H_{22}O_{10}$ | 446.1213 | 1.00 ± 0.13 (12.67%) |
| **7** | Chrysophanol-1-*O*-β-D-glucoside | 18.23 | 415.1030 | 0.2 | 277.0491, 253.0504, 225.0786 | $C_{21}H_{20}O_9$ | 416.1107 | 0.66 ± 0.10 (14.66%) |
| **8** | Physcion | 27.83 | 283.0605 | −0.4 | 240.0323 | $C_{16}H_{12}O_5$ | 284.0685 | 0.59 ± 0.08 (13.55%) |
| **9** | Rhein-8-*O*-β-D-glucoside | 13.76 | 445.0768 | −0.7 | 283.0244, 239.0344 | $C_{21}H_{18}O_{11}$ | 446.0849 | 0.40 ± 0.06 (15.60%) |
| **10** | Methylisorhein | 21.50 | 297.0392 | −2.4 | 253.0485 | $C_{16}H_{10}O_6$ | 298.0477 | 0.39 ± 0.03 (6.62%) |
| **11** | Aloe-emodin | 21.04 | 269.0457 | 2.6 | 240.03 64, 211.1301, 183.0977 | $C_{15}H_{10}O_5$ | 270.0528 | 0.31 ± 0.04 (12.29%) |
| **12** | Emodin-1-*O*-β-D-glucoside | 16.14 | 431.0965 | −3.0 | 269.0457, 239.0308 | $C_{21}H_{20}O_{10}$ | 432.1056 | 0.18 ± 0.03 (19.54%) |
| **13** | Aloe-emodin-8-*O*-(6'-*O*-acetyl)-glucoside isomer-2 | 21.04 | 473.1061 | −4.9 | 269.0444 | $C_{23}H_{22}O_{11}$ | 474.1162 | 0.14 ± 0.05 (38.73%) |
| **14** | Aloe-emodin-8-*O*-β-D-glucoside | 12.68 | 431.0977 | −0.2 | 269.0455 | $C_{21}H_{20}O_{10}$ | 432.1056 | 0.13 ± 0.02 (16.60%) |
| **15** | Methylisorhein-*O*-β-D-glucoside | 11.65 | 459.0927 | 0 | 297.0687 | $C_{22}H_{20}O_{11}$ | 460.1006 | 0.11 ± 0.01 (11.83%) |
| **16** | Aloe-emodin-8-*O*-(6'-*O*-acetyl)-glucoside isomer-3 | 19.78 | 473.1066 | −3.8 | 269.0431 | $C_{23}H_{22}O_{11}$ | 474.1162 | 0.10 ± 0.02 (19.83%) |
| **17** | Citreorosein isomer-1 | 16.96 | 285.0400 | 0.4 | 239.1045 | $C_{15}H_{10}O_6$ | 286.0477 | 0.10 ± 0.01 (13.93%) |
| **18** | 6-Methyl-aloe-emodin | 19.88 | 283.0600 | −2.1 | 240.1005 | $C_{16}H_{12}O_5$ | 284.0685 | 0.08 ± 0.01 (17.08%) |
| **19** | Citreorosein isomer-2 | 20.23 | 285.0401 | 0.7 | 239.0864 | $C_{15}H_{10}O_6$ | 286.0477 | 0.08 ± 0.01 (17.53%) |
| **20** | 1-Methyl-8-hydroxyl-9,10-anthraquinone-3-*O*-(6'-*O*-cinnamoyl)-glucoside | 24.73 | 589.1347 | 0.2 | 295.0577, 253.0498 | $C_{31}H_{26}O_{12}$ | 590.1424 | 0.06 ± 0.01 (9.26%) |
| **21** | Aloe-emodin-8-*O*-(6'-*O*-acetyl)-glucoside isomer-3 | 18.13 | 473.1054 | −6.3 | 269.0421 | $C_{23}H_{22}O_{11}$ | 474.1162 | 0.05 ± 0.01 (13.57%) |
| **22** | Isochrysophanol | 24.73 | 253.0498 | −1.2 | 225.0758 | $C_{15}H_{10}O_4$ | 254.0579 | 0.04 ± 0.01 (17.42%) |
| *Anthrones* | | | | | | | | |
| **31** | Cascaroside C | 13.57 | 563.1768 | 0.5 | 443.1341, 281.0717 | $C_{27}H_{32}O_{13}$ | 564.1843 | 0.16 ± 0.02 (14.63%) |
| **32** | Reidin B | 11.20 | 507.1121 | 8.1 | 313.0471, 169.0182 | $C_{30}H_{20}O_8$ | 508.1158 | 0.13 ± 0.05 (36.27%) |
| **33** | Reidin C | 13.21 | 537.1200 | 2.6 | 257.0790 | $C_{31}H_{22}O_9$ | 538.1264 | 0.08 ± 0.02 (27.51%) |
| **34** | Sennoside A | 15.41 | 861.1835 | −5.0 | 699.1339, 431.0891, 386.1003 | $C_{42}H_{38}O_{20}$ | 862.1956 | 0.06 ± 0.01 (18.05%) |

| | | | | | | | |
|---|---|---|---|---|---|---|---|
| *Galloylglucoses* | | | | | | | |
| **41** | 1-*O*-Galloyl-2-*O*-cinnamoyl-glucose isomer-1 | 15.28 | 461.1087 | 0.7 | 313.0550, 160.0123, 125.0238 | $C_{22}H_{22}O_{11}$ | 462.1162 | 0.76 ± 0.22 (28.9-%) |
| **42** | Gallic acid | 2.57 | 169.0137 | 0 | 125.0227, 78.9810 | $C_7H_6O_5$ | 170.0215 | 0.42 ± 0.04 (9.38%) |
| **43** | Gallic acid-3-*O*-glucoside | 2.43 | 331.0665 | 0 | 169.0120, 125.0216 | $C_{13}H_{16}O_{10}$ | 332.0743 | 0.40 ± 0.14 (33.98%) |
| **44** | 1,6-Di-*O*-galloyl-2-*O*-cinnamoyl-glucose isomer-1 | 16.16 | 613.1169 | −3.9 | 465.0664, 313.0551, 169.0165 | $C_{29}H_{26}O_{15}$ | 614.1272 | 0.39 ± 0.11 (26.99%) |
| **45** | 1-*O*-Galloyl-2-*O*-p-coumaroyl-glucose | 10.78 | 477.1032 | −0.2 | 313.0494 | $C_{22}H_{22}O_{12}$ | 478.1111 | 0.22 ± 0.09 (39.23%) |
| **46** | 1-*O*-Galloyl-2-*O*-cinnamoyl-glucose isomer-2 | 16.21 | 461.1086 | 0.4 | 160.9739 | $C_{22}H_{22}O_{11}$ | 462.1162 | 0.19 ± 0.04 (18.87%) |
| **47** | 1,6-Di-*O*-galloyl-2-*O*-cinnamoyl-glucose isomer-2 | 16.98 | 613.1206 | 2.1 | 169.0077 | $C_{29}H_{26}O_{15}$ | 614.1272 | 0.15 ± 0.03 (20.99%) |
| **48** | 6'-*O*-Galloylsucrose isomer-1 | 4.88 | 493.1183 | −2.0 | 313.0530, 169.0069 | $C_{19}H_{26}O_{15}$ | 494.1272 | 0.14 ± 0.03 (20.54%) |
| **49** | 6'-*O*-Galloylsucrose isomer-2 | 5.29 | 493.1197 | 0.8 | 313.0460, 169.0041 | $C_{19}H_{26}O_{15}$ | 494.1272 | 0.14 ± 0.02 (18.02%) |
| **50** | 6'-*O*-Galloylsucrose isomer-3 | 5.43 | 493.1197 | 0.8 | 313.0651 | $C_{19}H_{26}O_{15}$ | 494.1272 | 0.09 ± 0.01 (11.51%) |
| **51** | Gallic acid 3-*O*-(6'-*O*-galloyl)-glucoside isomer-1 | 18.24 | 483.0758 | −3.5 | 313.0829 | $C_{20}H_{20}O_{14}$ | 484.0853 | 0.09 ± 0.05 (55.34%) |
| **52** | Gallic acid 3-*O*-(6'-*O*-galloyl)-glucoside isomer-2 | 18.61 | 483.0795 | 4.1 | 313.0763 | $C_{20}H_{20}O_{14}$ | 484.0853 | 0.06 ± 0.03 (47.95%) |
| *Flavonoids* | | | | | | | | |
| **61** | (+)-Catechin | 7.65 | 289.0710 | −0.7 | 245.0821, 137.0237, 123.0429 | $C_{15}H_{14}O_6$ | 290.0790 | 0.56 ± 0.21 (36.87%) |
| **62** | (–)-Epicatechin-3-*O*-gallate | 11.34 | 441.0811 | −2.5 | 289.0714, 271.0578, 169.0135 | $C_{22}H_{18}O_{10}$ | 442.0900 | 0.10 ± 0.03 (31.96%) |
| **63** | Vitexin | 12.68 | 431.0974 | −0.9 | 311.0540, 283.0406, 116.9287 | $C_{21}H_{20}O_{10}$ | 432.1056 | 0.09 ± 0.02 (17.24%) |
| **64** | Eriodictyol | 15.41 | 287.0554 | −0.7 | 151.0034, 135.0441 | $C_{15}H_{12}O_6$ | 288.0634 | 0.05 ± 0.01 (19.09%) |
| *Naphthalenes* | | | | | | | | |
| **71** | Torachrysone-8-*O*-β-D-glucoside | 17.48 | 407.1343 | 0.2 | 245.0813, 230.0487 | $C_{20}H_{24}O_9$ | 408.1420 | 0.38 ± 0.06 (17.20%) |
| **72** | Rheumone A | 17.01 | 417.1186 | 0 | 270.0633 | $C_{21}H_{22}O_9$ | 418.1264 | 0.14 ± 0.02 (14.56%) |
| **73** | 6-Hydroxy-musizin-8-*O*-β-D-glucoside | 14.90 | 393.1180 | −1.5 | 231.0643 | $C_{19}H_{22}O_9$ | 394.1264 | 0.09 ± 0.03 (31.78%) |
| *Stilbenes* | | | | | | | | |
| **81** | Resveratrol-4'-*O*-β-D-glucoside | 10.53 | 389.1230 | −1.5 | 227.0996 | $C_{20}H_{22}O_8$ | 390.1315 | 0.08 ± 0.02 (29.80%) |
| **82** | Rhapontigenin | 15.49 | 257.0814 | 0 | 242.0582, 241.0483, 213.0457 | $C_{15}H_{14}O_4$ | 258.0892 | 0.05 ± 0.01 (24.29%) |
| **83** | cis-3,4',5-Trihydroxystlbene-4'-*O*-(2''-*O*-galloyl)-glucoside | 13.41 | 541.1345 | −0.2 | 313.0644, 125.0320 | $C_{27}H_{26}O_{12}$ | 542.1424 | 0.04 ± 0.01 (24.71%) |
| **84** | Resveratrol | 13.96 | 227.0711 | 1.3 | 185.0599, 143.0526 | $C_{14}H_{12}O_3$ | 228.0786 | 0.02 ± 0.00 (19.15%) |
| *Pyranones* | | | | | | | | |
| **91** | 2,5-Dimethyl-7-hydroxychromone | 15.05 | 189.0552 | 0 | 174.0294, 146.0354 | $C_{11}H_{10}O_3$ | 190.0630 | 0.05 ± 0.01 (10.19%) |
| **92** | 2-Methyl-5-acetonyl-7-hydroxychromone | 14.92 | 231.0658 | 0.4 | 189.0547, 188.0492 | $C_{13}H_{12}O_4$ | 232.0736 | 0.05 ± 0.02 (35.39%) |
| **93** | 2-(2'-Hydroxypropyl)-5-methyl-7-hydroxychrom | 14.48 | 233.0813 | −0.4 | 189.0569 | $C_{13}H_{14}O_4$ | 234.0892 | 0.03 ± 0.00 (11.55%) |

| | | | | | | | | |
|---|---|---|---|---|---|---|---|---|
| | one isomer-1 | | | | | | | |
| **94** | 2-(2'-Hydroxypropyl)-5-methyl-7-hydroxychromone isomer-2 | 16.33 | 233.0806 | 3.4 | 189.0361 | $C_{13}H_{14}O_4$ | 234.0892 | 0.01 ± 0.00 (18.27%) |
| *Phenylbutanones* | | | | | | | | |
| **101** | Lindleyin or Isolindleyin | 12.08 | 477.1390 | −1.5 | 313.0547, 169.0131, 123.0130 | $C_{23}H_{26}O_{11}$ | 478.1475 | 0.09 ± 0.01 (13.69%) |
| **102** | 4-(4'-Hydtoxyphenyl)-2-butanone-4'-*O*-(2"-*O*-galloyl-6"-*O*-cinnamoyl)-glucoside | 20.63 | 607.1804 | −2.0 | 443.0992 | $C_{32}H_{32}O_{12}$ | 608.1894 | 0.03 ± 0.01 (16.43%) |

The dose level data represent the mean ± standard deviation for samples of 14 lots of LianhuaQingwen.

**Table S4** Unbound fractions of Dahuang compounds in human plasma ($f_{u\text{-plasma}}$).

| Compound (ID) | $f_{u\text{-plasma}}$ |
|---|---|
| Rhein (**2**) | 0.0028 ± 0.0001 |
| Methylisorhein (**10**) | 0.0019 ± 0.0001 |
| 4-*O*-Methylgallic acid (**M42$_{M2}$**) | 0.42 ± 0.02 |

**Table S5** Estimation of maximum absorbable dose (MAD) of Dahuang anthraquinones.

| Compound (ID) | Equilibrium solubility[a] (μg/mL) | MAD[b] (μg/person) | Daily dose (μg/person) |
|---|---|---|---|
| Chrysophanol (**1**) | < 0.05 | < 10.1 | 554.9 |
| Emodin-8-*O*-β-D-glucoside (**2**) | < 0.1 | < 20.3 | 444.4 |
| Rhein (**3**) | ≤ 0.1 | ≤ 20.3 | 576.2 |
| Chrysophanol-8-*O*-β-D-glucoside (**4**) | 3.49 | 706.7 | 624.8 |
| Emodin (**5**) | < 0.05 | < 101.3 | 314.7 |
| Physcion-8-*O*-β-D-glucoside (**6**) | < 0.05 | < 10.1 | 510.6 |
| Chrysophanol-1-*O*-β-D-glucoside (**7**) | 3.22 | 652.1 | 202.2 |
| Physcion (**8**) | < 0.05 | < 10.1 | 213.2 |
| Rhein-8-*O*-β-D-glucoside (**9**) | 0.63 | 127.6 | 129.9 |
| Methylisorhein (**10**) | ≤ 0.1 | ≤ 20.3 | 175.2 |
| Aloe-emodin (**11**) | < 0.05 | < 101.3 | 83.4 |

[a] Solubility was measured at 37°C by slurrying the solid Dahuang anthraquinone substance in deionized water for 48 h, passing the slurry through a 0.22 μm filter, and analyzing the filtrate using UPLC-UV spectrometry.

[b] Maximum absorbable dose (MAD) was estimated using the following equation: MAD = S × $K_a$ × SIWV × SITT, where S is solubility at pH 7.0, $K_a$ is transintestinal absorption rate constant, SIWV is small intestinal water volume (assumed to be ~250 mL), SITT is small intestinal transit time (assumed to be ~270 min) [1]. 0.003 $min^{-1}$ is a typical low $K_a$ for Dahuang with Caco-2-based low to intermediate membrane permeability (i.e., **1**, **2**, **3**, **4**, **6**, **7**, **8**, **9**, and **10**), while the higher value of 0.03 $min^{-1}$ is typical of Dahuang anthraquinones with the high membrane permeability (i.e., **5** and **11**).

**Table S6** Net transport ratios for *in vitro* transport of rhein (**3**) and methylisorhein (**10**) mediated by various human efflux transporters.

| Compound (ID) | Net transport ratio | | | | | |
| --- | --- | --- | --- | --- | --- | --- |
| | MDR1 | BCRP | MRP1 | MRP2 | MRP3 | MRP4 |
| Positive substrate | 13.8 ± 1.7 | 11.4 ± 0.8 | 12.2 ± 2.0 | 7.1 ± 1.3 | 15.3 ± 0.5 | 11.6 ± 1.8 |
| Rhein (**3**) | 2.3 ± 0.5 | 2.7 ± 0.1 | 1.6 ± 0.6 | 3.1 ± 0.3 | 2.4 ± 0.7 | 2.0 ± 0.4 |
| Methylisorhein (**10**) | 3.5 ± 0.1 | 2.9 ± 0.2 | 3.4 ± 0.3 | 3.4 ± 0.4 | 3.4 ± 0.7 | 3.0 ± 0.4 |

For assessment of vesicular uptake mediated by human efflux transporters, differential transport rates between transfected vesicles and mock vesicles were defined as a net transport ratio, which represents the mean ± standard deviation (n = 3). A net transport ratio > 2 suggested a positive result. Positive substrates were glycyrrhizin (for MDR1 and BCRP) and estradiol-17β-D-glucuronide (MRP1, MRP2, MRP3, and MRP4), and their final concentrations were 10 μmol/L with incubation time of 10 min.

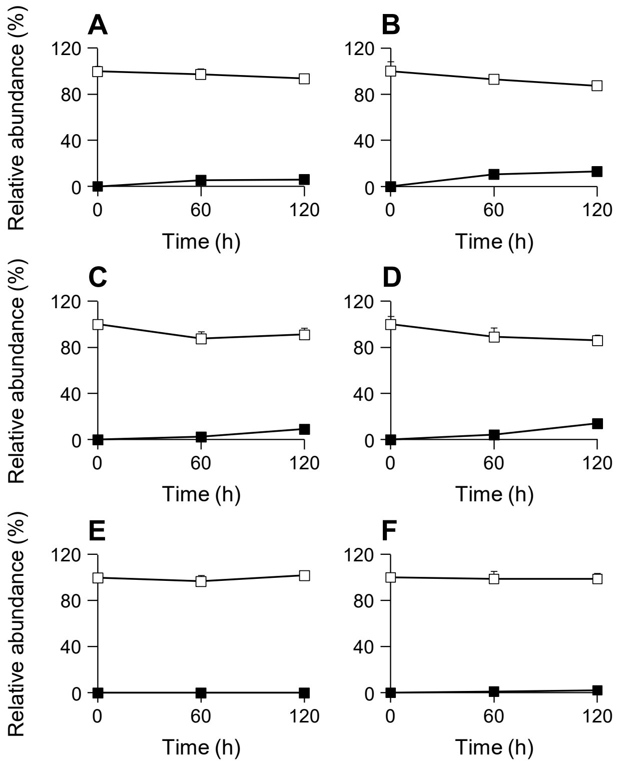


**Fig. S1.** *In vitro* metabolism of rhein (**3**) and methylisorhein (**10**) mediated by human enzymes. Glucuronidation of rhein (A) and methylisorhein (B); sulfation of rhein (C) and methylisorhein (D); oxidation of rhein (E) and methylisorhein (F). Solid dots, rhein or methylisorhein; open dots, the respective metabolites.

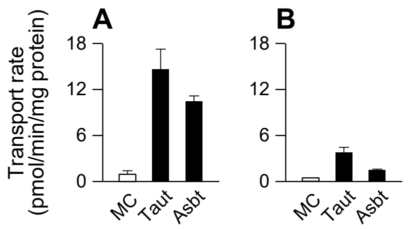


**Fig. S2.** *In vitro* transport of rhein (**3**) and methylisorhein (**10**) mediated by rat transporters. (A) Transport of rhein by rat Taut- and Asbt-transfected HEK-293 cells. (B) Transport of methylisorhein by rat Taut- and Asbt-transfected HEK-293 cells. MC, mock HEK-293 cells.

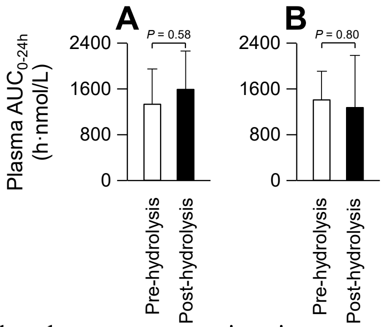


**Fig. S3.** Comparison of area under plasma concentration-time curve (AUC) of free and total rhein (**3**, A) or methylisorhein (**10**, B) in humans after oral administration of LianhuaQingwen at 12 capsules/person. The plasma samples underwent pre- and post-hydrolysis treatments with hydrochloric acid (4 M) at 80°C for 30 min to release free anthraquinone from its various forms, including glycosylated, glucuronidated, and sulfated forms.

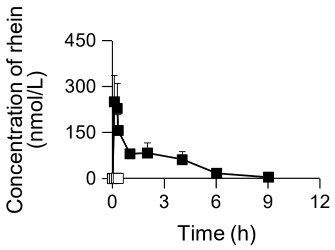


**Fig. S4.** Plasma concentration-time profiles of rhein (**3**) and rhein-8-*O*-β-D-glucoside (**9**) in rats after oral administration of rhein-8-*O*-β-D-glucoside at 1.8 mg/kg. Solid dots, rhein; Open dots, rhein-8-*O*-β-D-glucoside.
